# Outbreak and Postnatal Antibiotic Exposures Drive the Development Trajectory of the Nasopharyngeal Microbiota in the First Year of Life

**DOI:** 10.1101/2023.09.14.23295567

**Authors:** Polona Rajar, Achal Dhariwal, Gabriela Salvadori, Heidi Aarø Åmdal, Dag Berild, Ulf R. Dahle, Drude Fugelseth, Gorm Greisen, Ulrik Lausten-Thomsen, Ola Didrik Saugstad, Fernanda Cristina Petersen, Kirsti Haaland

**Affiliations:** Department of Neonatal Intensive Care, Division of Paediatric and Adolescent Medicine, Oslo University Hospital, Oslo, Norway; Institute of Oral Biology, Faculty of Dentistry, University of Oslo, Oslo, Norway; Department of Infectious Diseases, Oslo University Hospital, Oslo, Norway; Institute of Clinical Medicine, Faculty of Medicine, Oslo University, Oslo, Norway; Centre for Antimicrobial Resistance, Norwegian Institute of Public Health, Oslo, Norway; Department of Neonatology, Copenhagen University Hospital Rigshospitalet, Copenhagen, Denmark; Department of Pediatric Research, University of Oslo, Oslo, Norway

**Keywords:** whole metagenomic sequencing, nasopharyngeal, respiratory, microbiome, preterm birth, hospital outbreak, antibiotics

## Abstract

Early exposure to antibiotics and prolonged hospitalization in preterm infants may perturb microbiome development and contribute to adverse health outcomes. Although nasopharyngeal microbiomes are linked to respiratory infections, their early development is underexplored and often assessed with 16S rRNA sequencing, which lacks species-level resolution. Here, we investigated nasopharyngeal microbiota dynamics in 66 preterm infants by performing whole metagenome sequencing on 345 nasopharyngeal aspirates collected from birth until 6 months corrected age (∼7-10 months chronological age). The nasopharyngeal microbiota exhibited highly individualized and age-structured patterns shaped by postnatal antibiotic exposure and hospitalization. Postnatal antibiotic exposure had pronounced effects on microbial diversity, composition, stability and maturation, with some effects persisting to six months corrected age. An unexpected *Serratia marcescens* outbreak in the NICU left a lasting signature, with persistent carriage observed in over 90% of post-outbreak infants at 6 months corrected age. *S. marcescens* established a stable, self-reinforcing community type emerging exclusively after the outbreak, with a machine learning model accurately predicting persistent carriage from early microbiome composition, even after excluding *S. marcescens* from the analysis. These findings underscore the importance of understanding how postnatal antibiotics and NICU outbreaks shape nasopharyngeal microbiome trajectories, with potential long-term respiratory health implications.

## INTRODUCTION

Microbial colonization after birth is a dynamic process, influenced by environmental and host factors, which holds significant implications for long-term health ^1–3^. Most of the current knowledge originates from studies on the intestinal microbiota, leaving a knowledge gap regarding the microbiota development in other body niches, particularly in the respiratory tract, where infections are notably common and sometimes critical ^4–6^.

Immediately after birth, the upper respiratory tract mucosa is rapidly colonized by microbes originating from mothers’ genito-rectal, skin and oral flora, as well as from the surrounding environment. This initial colonization is followed by an increase in bacterial abundance and a shift towards distinct bacterial communities within weeks ^3, 7, 8^. For term infants, the nasopharyngeal microbiota further differentiates towards stable microbial profiles by six months of age, influenced by factors such as the mode of delivery, nutrition regime, and antibiotic therapy ^3, 6, 7, 9^. Signals from the respiratory microbiota are necessary for the infant’s immune system maturation ^10–13^. Alterations in airway microbiota during this critical developmental phase have been linked to severity and susceptibility to respiratory diseases later in life, such as asthma, otitis media, and respiratory syncytial virus infections ^14–19^.

Preterm infants are particularly vulnerable due to the development of inflammation and infection-related pathologies in the respiratory tract, even without confirmed neonatal lung injury ^20–23^. For preterm infants, disruption in microbiota development may pose additional risks, exacerbated by prolonged hospitalization and early antibiotic treatment ^2, 24^. Both factors dramatically influence the microbial ecological succession in preterm infants, though their effects on non-intestinal sites remain less understood. In Norway, 77% of infants born at gestational age (GA) <32 weeks are exposed to antibiotics during their first week of life, and 75% are treated with antibiotics within 72 hours after birth due to suspected early onset neonatal sepsis (EONS) ^25^.

To date, research on the respiratory microbiome has primarily utilized 16S rRNA amplicon sequencing. This method, however, lacks the species-level resolution necessary to distinguish between commensals and potential pathogens ^20^. This gap in knowledge is largely attributed to the challenges associated with low microbial biomass and the high content of human DNA in nasopharyngeal samples, which complicates the retrieval of sufficient microbial DNA for non-targeted sequencing using shotgun metagenomics ^26^. To address these challenges, we recently developed and optimized a sample processing workflow combining host DNA depletion (MolYsis™) with lytic extraction (MasterPure™), enabling whole-metagenome sequencing (WMS) of nasopharyngeal aspirates from preterm infants for comprehensive species-level taxonomic profiling and resistome characterization ^27, 28^. Here, we focus on the developmental dynamics of the nasopharyngeal microbiome in preterm infants and explore the impact of different clinical variables and exposures on its trajectories and maturation.

## MATERIALS & METHODS

### Study design and sample collection

Between July 2019 and January 2021, we approached parents within 48 hours after birth of all infants with a gestational age from 28 weeks 0 days to 31 weeks 6 days who were born at or transferred to the Ullevål Neonatal Intensive Care Unit (NICU), a tertiary NICU at Oslo University Hospital. Patients’ metadata were collected from the electronic records. We divided infants into three groups (Table 1): “Only Early Antibiotics” (*n* = 24), “Other Antibiotics” (*n* = 21) and “Naive” (*n* = 21). For brevity, these groups are referred to as “Only Early AB”, “Other AB” and “Naive” in figures and tables throughout. Sample metadata are provided in Supplementary Table E1. Infants in the “Only Early Antibiotics” group were treated with intravenous ampicillin and gentamicin due to suspected early-onset neonatal sepsis (EONS), initiated within 24 hours after birth and discontinued after a mean (SD) duration of 4 (2) days. The “Other Antibiotics” group received different combinations of either early antibiotics (initiated within 72 hours after birth); late antibiotics (initiated later than 72 hours after birth); and/or antibiotics after discharge (intravenous or per oral route). Five infants received antibiotics in the period between discharge from the NICU and six months corrected age. The “Naive” group included infants who did not receive any antibiotics from birth until six months corrected age.

**Table 1.** Cohort demographics. AB: antibiotic; GA: gestational age; BW: birth weight; LBW: low birth weight (from 1500g to < 2500g); VLBW: very low body weight (from 1000g to < 1500g); ELBW: extremely low body weight (< 1000g); ROM: rupture of membranes; MOM: mothers’ own milk. * *p*<0.05, ** *p*<0.001.

| Whole cohort<br>(n=66) | AB naive<br>(n=21) | AB exposed<br>(n=45) | Only early ABs<br>(n=24) | Other ABs<br>(n=21) |
| --- | --- | --- | --- | --- |
| Male, n (%) | 12 (57%) | 33 (73%) | 18 (75%) | 15 (71%) |
| GA, weeks (mean,<br>SD) | 31, 6/7 | **29 4/7, 1 \*\*** | **29 4/7, 1 \*\*** | **29 3/7, 1 \*\*** |
| BW, grams (mean,<br>SD) | 1562, 188 | **1280, 262 \*\*** | **1278, 255 \*\*** | **1282, 275 \*\*** |
| BW, group, n (%) |  | * | * | * |
| ELBW | 0 (0%) | **6 (13%)** | **3 (12%)** | **3 (14%)** |
| VLBW | 8 (38%) | **31 (69%)** | **17 (71%)** | **14 (67%)** |
| LBW | 13 (62%) | **8 (18%)** | **4 (17%)** | **4 (19%)** |
| Vaginal delivery, n<br>(%) | 4 (19%) | 16 (35.5%) | 9 (37.5%) | 8 (38%) |
| Caesarian section,<br>n (%) | 17 (81%) | 29 (64.5%) | 15 (62.5%) | 13 (62%) |
| ABs during<br>pregnancy, n (%) |  |  |  |  |
| None | 21 (100%) | **32 (71%) \*** | **13 (54%) \*\*** | 19 (90%) |
| < 10 days | 0 (0%) | 10 (22%) | 10 (42%) | 0 (0%) |
| ≥10days | 0 (0%) | 3 (7%) | 1 (4%) | 2 (10%) |
| Intrapartum AB<br>prophylaxis, n (%) |  |  |  |  |
| None documented | 2 (10%) | 3 (7%) | 1 (4%) | 2 (10%) |
| Given | 19 (90%) | 42 (93%) | 23 (96%) | 19 (90%) |
| ROM, hours<br>(means, SD) | 3, 12 | 201, 491 | **258, 422 \*** | 136, 563 |
| Apgar at 10 min<br>(mean, SD) | 9, 1 | 9, 1 | 8, 1 | 9, 1 |
| Nutrition, n (%) |  |  |  |  |
| Fully breastfed | 7 (39%) | 13 (30%) | 5 (22%) | 8 (40%) |
| > 50% MOM | 3 (17%) | 11 (26%) | 7 (30%) | 4 (20%) |
| > 50% formula | 6 (33%) | 8 (18%) | 5(22%) | 3(15%) |
| Only formula | 2 (11%) | 11 (26%) | 6 (26%) | 5 (25%) |
| Lost to follow up | 3 (14%) | 2 (4%) | 1 (4%) | 1(5%) |

Nasopharyngeal aspirates (NPAs) were obtained within 48 hours of six predefined time points: day of life (DOL) 0 (day of birth; time point 1), DOL 7 (time point 2), DOL 14 (time point 3), DOL 28 (time point 4), DOL 56 or at NICU discharge – whichever came first (time point 5) and at six months corrected age (time point 6), where DOL is defined as the number of days elapsed since birth. Additional samples were obtained within 48 hours of antibiotic initiation and discontinuation, which in most cases coincided with the predefined time points (Figure S1A, B). If collection was not possible at the designated time due to clinical instability, it was postponed until the infant was stable. Briefly, a sterile suction catheter was inserted along the nasal wall into the nasopharynx, vacuum suction was applied for 5 seconds, and 2 ml of sterile 20% glycerol solution was immediately suctioned through the catheter for sample preservation. Full details of the collection protocol have been described previously ^28^. Samples were stored at −80°C in 20% glycerol (2 ml) solution until further processing in the laboratory. Throughout all further steps, samples belonging to the same infant were processed concomitantly.

### Metagenomic DNA extraction, library preparation and sequencing

Metagenomic DNA extraction was performed using a previously optimized workflow combining host DNA depletion (MolYsis™ Basic5, Molzym) with lytic extraction (MasterPure™ Gram Positive DNA Purification Kit, Epicentre), specifically developed for low-biomass nasopharyngeal samples from preterm infants ^28^, with minor modifications detailed in Supplementary Methods. All samples were spiked with 20 µl of Spike-in Control II Low Microbial Load (Catalog D6321 & D6321-10, ZymoBIOMICS™, Irvine, CA), prior to DNA extraction.

Metagenomic libraries were prepared using the Nextera DNA Flex kit (Illumina Inc., San Diego, CA) with twelve PCR amplification cycles as previously optimized for low-input DNA samples ^28^; full library preparation details are provided in the Supplementary Methods. Whole metagenome sequencing (WMS) was conducted at the Norwegian Sequencing Centre (Oslo, Norway), on a NovaSeq S4 platform (Illumina Inc.) using a paired-end sequencing approach with a targeted read length of 150 bp in high-output mode.

For downstream analysis, nine samples with zero read counts after contaminant removal were excluded. Additionally, for four infants with more than one sample collected at the same time point, one sample per time point was retained, excluding five samples in total. Details on sample inclusion and exclusion at each processing stage are provided in Supplementary Table E2.

### Bioinformatics processing and microbiome profiling

FASTQC (v.0.11.9) was used to assess the quality of raw and clean reads ^29^. Quality and adapter trimming were performed with Trim Galore (v.0.6.1) ^30^ with default parameters. Quality-filtered reads were aligned to the human reference genome (GRCh38) and human DNA sequences were removed using Bowtie2 (v.2.3.4.2; *-N 1 -k 1 --fr --end-to-end --phred33 --very-sensitive --no-discordant*) ^31^. MetaPhlAn 4 (v.4.2.4) ^32^ was used for taxonomic profiling of the remaining high-quality non-host reads using the mpa_vJan25_CHOCOPhlAnSGB_202503 database, which includes clade-specific marker genes for mock community spike-in species, with default parameters.

### Statistical analysis and data visualization

Statistical analysis of cohort demographics was performed in STATA SE 17 using unpaired t-test, Pearson’s Chi-squared, Fisher’s exact test, and one-way ANOVA. All downstream WMS data analyses and 16S rRNA qPCR statistical analyses were conducted in R (v.4.2.1) within RStudio (v.2022.07.2+576) ^33, 34^. Figures were generated primarily using the ggplot2 (v.3.4.0) package ^35^, with some figures generated using and GraphPad Prism 9.4.1(458)© GraphPad Software, LLC (Boston, MA, USA). For independent between-group comparisons, non-parametric Wilcoxon rank-sum tests or Kruskal–Wallis tests with pairwise Wilcoxon rank-sum post-hoc tests were applied depending on data type and distribution. Spearman correlation was used to assess associations between continuous variables. A *p*-value < 0.05 was considered statistically significant throughout, unless otherwise stated. *P*-values were corrected for multiple testing using the Benjamini–Hochberg (BH) method where appropriate. Detailed descriptions of all downstream statistical analyses and bioinformatic methods are provided in the Supplementary Methods.

## RESULTS

### Study cohort characteristics and overview of sequencing results

We obtained 369 nasopharyngeal samples from 66 infants at six sampling time points between birth and six months corrected age (Figure 1), including 108 samples from 21 infants not exposed to any postnatal antibiotics (“Naive” group, Figure S1A, B). Infants exposed to postnatal antibiotics (*n* = 45) had lower GA and birth weight (BW), and their mothers were more often treated with antibiotics during pregnancy and had a more prolonged period of ruptured membranes before delivery compared to “Naive” infants (Table 1). The average (SD) early antibiotic (ampicillin and gentamicin) treatment duration was 4 (2) days. Blood cultures were always obtained before antibiotic administration; none of the infants had a positive blood culture prior to initiation of early antibiotics, and 13 had a positive blood culture consistent with late-onset sepsis (LOS) prior to initiation of late antibiotics.

**Figure 1.**
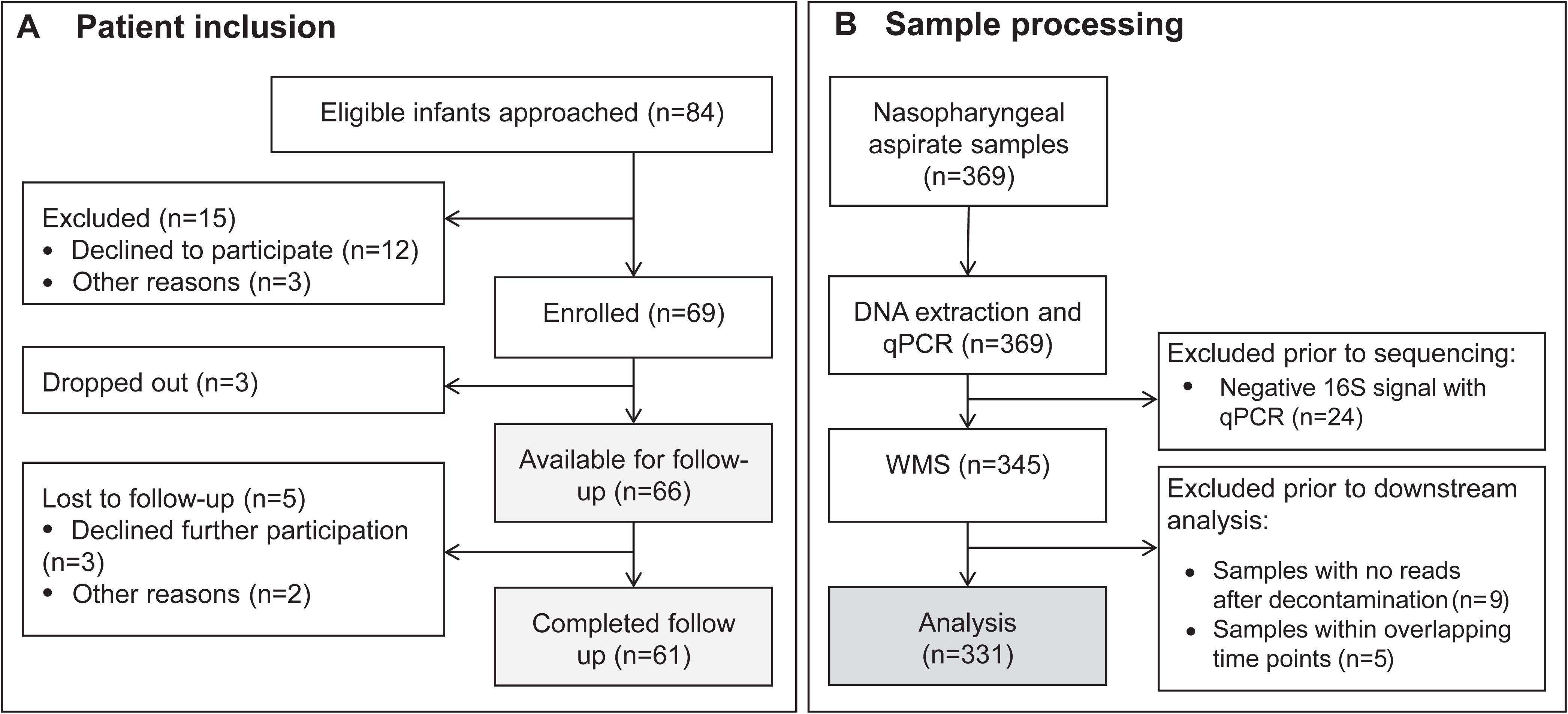
Patient inclusion and sample processing. (**A**) Flowchart describing patient inclusion process and availability of infants from birth until last follow up at six months corrected age. (**B**) Overview of obtained nasopharyngeal aspirate samples and excluded samples prior to main microbiome analysis. WMS: whole metagenome sequencing.

In total, 12.13 billion raw sequencing reads were generated from nasopharyngeal samples, with a median of 31.97 million (M) reads per sample (mean: 35.17 M; IQR: 27.72–39.67 M) (Supplementary Table E3). Of the total reads, 99.09–99.99% survived quality filtering and trimming across all samples. In contrast, the median number of raw sequencing reads detected in negative control samples was ∼2.5 logs lower (median: 0.09 M; mean: 0.15 M; IQR: 0.04– 0.27 M) compared to nasopharyngeal samples (Supplementary Table E3). Negative controls showed no structured microbial community profile, suggesting that amplification did not introduce any systematic compositional bias. On average, 55% of quality-filtered reads (range: 0.86%–98.6%) were associated with the human genome. Following removal of human reads, MetaPhlAn4 estimated a median bacterial content of 17.55 M reads per nasopharyngeal sample (mean: 21.78 M, IQR: 3.23–35.31 M; Supplementary Table E3), derived from clade-specific marker gene coverage. A total of 28 microbial species, including three ZymoBIOMICS spike-in species, were identified as contaminants and excluded from the raw microbial abundance tables (Supplementary Table E4). Of the 38 total samples excluded prior to downstream analysis, 31 belonged to antibiotic-exposed infants (Fisher’s exact test: *p* = 0.10; Supplementary Table E2). Overall, 331 nasopharyngeal samples collected from 66 preterm infants were included in downstream analysis, containing 384 bacterial species distributed across 13 bacterial phyla.

### Characterization of nasopharyngeal microbial load, composition, and dynamics in preterm infants

Firstly, we quantified total absolute bacterial abundance in nasopharyngeal samples from preterm infants using 16S rRNA gene-targeted qPCR. Using a linear mixed-effects model (LMM) with infant ID as random effect, we observed a significant increase in absolute bacterial abundance from birth (time point 1) through the first two months of life, reaching a plateau at time points 3–5 (estimated marginal means (emmeans): all *p.adj* > 0.13), followed by a significant decline at six months corrected age (time point 6 vs time points 3–5; emmeans: all *p.adj* < 0.05; Figure 2A). The mean absolute bacterial abundance across infant groups was lowest at time point 1 (mean log 16S qPCR: -1.23 ng), indicating low bacterial loads in the nasopharynx of preterm infants at birth (Figure 2B). Further, infants in the “Other Antibiotics” group had significantly lower bacterial loads at time point 3 compared to those in the “Only Early Antibiotics” group (*p.adj* = 0.001) and the Naive group (*p.adj* = 0.008; Figure 2B).

**Figure 2.**
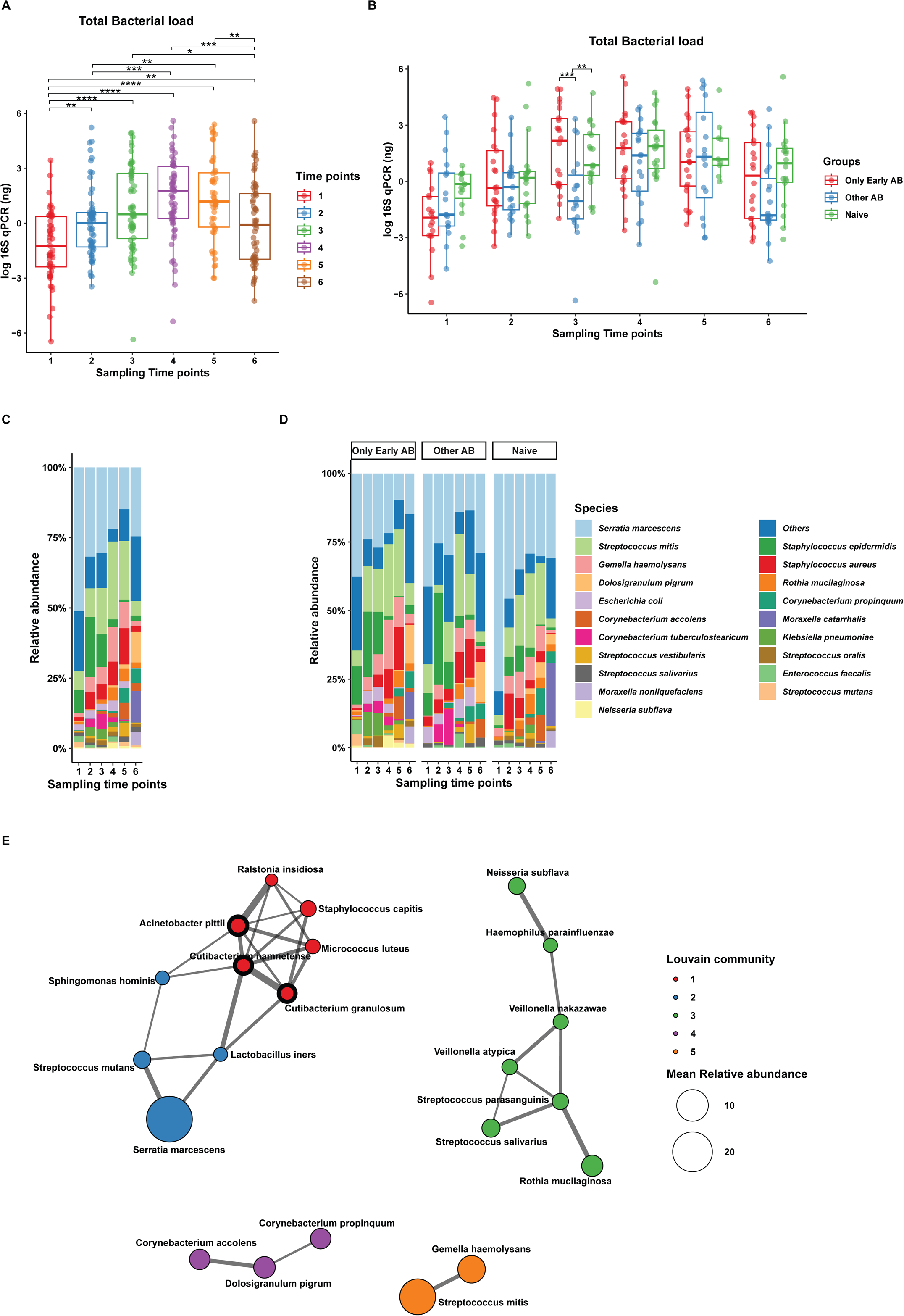
Taxonomic composition of the nasopharyngeal microbiome. **(A)** Boxplot showing the total bacterial DNA (log 16S rRNA gene qPCR, ng), for all infants across sampling time points spanning from birth to six months corrected age. **(B)** Comparison of total bacterial load stratified by postnatal antibiotic exposure group at each time point. In panels **(A)** and **(B)**, the central horizontal line in each boxplot represents the median, the box indicates the interquartile range (IQR), and the whiskers extend to 1.5× IQR. Significant differences were assessed using a linear mixed-effects model (LMM) with pairwise comparisons derived from estimated marginal means (emmeans) and BH correction (\**p* < 0.05, \*\**p* < 0.01, \*\*\**p* < 0.001, and \*\*\*\**p* < 0.0001). **(C–D)** Stacked bar plots displaying the mean relative bacterial abundance of the 20 most abundant taxa across sampling time points and AB exposure groups at the species level. Panels **(C)** illustrates the mean relative bacterial abundance by sampling time point (x-axis), while panels **(D)** presents the data stratified by both sampling time point and postnatal antibiotic exposure groups. Low abundant taxa are grouped into the “Others” category. **(E)** Microbial co-occurrence network inferred using SparCC correlations (1,000 bootstraps; *p.adj* < 0.05, |r| > 0.3), comprising 22 taxa and 32 edges, resolving into five subcommunities (density = 0.139; modularity Q = 0.557). Network visualized with Fruchterman–Reingold layout, with nodes colored by Louvain community and sized by mean relative abundance. Hub taxa (top 90th percentile by degree) are outlined in bold.

We next characterized the microbial community composition in the developing nasopharyngeal microbiome using shotgun metagenomic sequencing. Absolute bacterial abundance (measured by qPCR) was moderately correlated with the estimated bacterial genome-derived read counts (Spearman: ρ = 0.50, *p* < 0.001; Figure S1C), suggesting that sequencing-derived read counts partially reflect underlying bacterial load. Firmicutes (median: 44.3%; IQR: 5.7–89.5%) and Proteobacteria (median: 17.9%; IQR: 0.9–89.8%) were the dominant phyla across all groups and time points in the nasopharyngeal microbiota, followed by Actinobacteria (median: 0.8%; IQR: 0.01–11%) (Figure S2A, B).

At the genus level, the five most abundant genera were *Serratia* (mean: 29.1%; median: 0.5%; IQR: 0-71.6%), *Streptococcus* (mean: 20.5%; median: 5.6%; IQR: 0.1-34.3%), *Staphylococcus* (mean: 15.2%; median: 5.6%; IQR: 0.1–15.1%), *Corynebacterium* (mean: 6.6%) and *Gemella* (mean: 6.2%) (Figure S2C). *Staphylococcus* and *Streptococcus* were the most prevalent genera, detected in over 85% of samples. *Serratia* predominated in the “Naive” group (mean: 41.4%) and “Other Antibiotics” group (mean: 25.6%), while *Streptococcus* predominated in the “Only Early Antibiotics” group (mean: 23.2%), consistent with the predominantly post-outbreak recruitment of Naive infants (Figure S2D). At time point 1, *Serratia* had the highest mean relative abundance (51.1%), followed by *Streptococcus* (12.3%) and *Staphylococcus* (11.1%).

*Streptococcus mitis* and *Staphylococcus epidermidis* were the most prevalent species, identified in over 70% of samples. The most abundant species were *Serratia marcescens* (mean: 29.1%; median: 0.5%; IQR: 0-60.2%), reflecting its outbreak-driven variable distribution across samples, followed by *S. mitis* (mean: 14.5%; median: 1.6%; IQR: 0– 18.6%), *S. epidermidis* (mean: 8.1%; median: 0.3%; IQR: 0–3.7%), *Gemella haemolysans* (mean: 6.2%; median: 0.01%; IQR: 0–2.1%), and *Staphylococcus aureus* (mean: 5.8%; median: 0.03%; IQR: 0–1.7%) (Figure 2C). *S. epidermidis* was dominant from birth, peaking at time point 2 (mean: 21.3%; IQR: 0.6–15.4%), before declining from time point 3 onwards, after which *S. aureus* became the dominant *Staphylococcus* species from time point 4 onwards (mean: 8.4–12.9%). *S. mitis* remained the most abundant *Streptococcus* species across all time points (Figure 2C). At six months corrected age (time point 6), *Dolosigranulum pigrum* was detected at comparable prevalence across groups (Only Early Antibiotics: 60%; Other Antibiotics: 47%; Naive: 61%), though mean relative abundance was higher in antibiotic-exposed groups (14.1% and 14.5% respectively) than in the Naive group (3.9%), where persistent high *S. marcescens* abundance (mean: 30.7%) likely suppressed other taxa through compositional effects. *Moraxella catarrhalis* showed higher mean abundance in the Naive group (23.1%) compared to antibiotic-exposed groups (Only Early Antibiotics: 9.7%; Other Antibiotics: 1.6%), though median abundance was 0% in all groups and prevalence remained low (15–39%), suggesting that this pattern was driven by a small subset of samples with high *Moraxella catarrhalis* abundance rather than consistent colonization across the group. Differential abundance analysis did not reveal statistically significant differences in either taxon between groups (ANCOM-BC2: all *p.adj* > 0.05; Figure 2D; Supplementary Table E5).

To further explore the interactions within the nasopharyngeal microbiota, we generated a co-occurrence network using SparCC (|r| > 0.3, *p.adj* < 0.05; Figure 2E). This comprised 22 taxa and 32 edges, resolving into five subcommunities. Hub taxa identified by eigenvector centrality included *Cutibacterium namnetense*, *Acinetobacter pittii*, and *Cutibacterium granulosum*, anchoring a subcommunity of skin-associated and opportunistic organisms. Notably, despite being the most abundant species, *Serratia marcescens* exhibited only two co-occurrence edges, occupying a peripheral subcommunity isolated from the broader network. Health-associated commensals *D. pigrum*, *Corynebacterium accolens*, and *Corynebacterium propinquum* formed a distinct subcommunity with no connections to *S. marcescens*, while oral-respiratory taxa including *Rothia mucilaginosa*, *Veillonella* spp., *Streptococcus parasanguinis*, and *Haemophilus parainfluenzae* comprised a fourth subcommunity. *S. mitis* and *G. haemolysans*, phylogenetically related commensals of the upper respiratory tract, co-occurred exclusively in a fifth subcommunity.

### Postnatal antibiotic exposure affects the nasopharyngeal microbiome diversity, composition and stability

Metagenomic analysis revealed an increase in species-level α-diversity (Shannon index) over time, with significant differences observed from time point 4 onwards compared to earlier time points (emmeans: all *p.adj* < 0.05; Figure 3A). To determine which clinical variables were associated with microbiota diversity (Shannon), we used an LMM approach including subject as a random effect to account for individual variation, with the significance of each variable assessed using likelihood ratio test (LRT). No clinical variable, other than postnatal age, was significantly associated with infant nasopharyngeal microbiome diversity (all *p.adj* > 0.05; Supplementary Table E6). In addition, sequencing depth had minimal influence on α-diversity estimates (Spearman: ρ = −0.215; Figure S9A). When stratified by postnatal antibiotic exposure, the nasopharyngeal microbiota followed distinct α-diversity trajectories across groups (time point × antibiotic exposure group interaction (LRT: *p* = 0.01; Figure 3B)). In the “Only Early Antibiotics” group, we observed a decline in α-diversity at time points 2 and 3, coinciding with the discontinuation of ampicillin and gentamicin, before recovering significantly and continuing to increase through six months corrected age (time point 2 vs 4: *p.adj* = 0.05; time point 2 vs 6: *p.adj* < 0.0001; Figure 3B). No significant differences between any time points were observed in the “Other Antibiotics” or “Naive” groups (all *p.adj* > 0.05). Notably, at six months corrected age, the “Only Early Antibiotics” group exhibited significantly higher α-diversity than both the “Other Antibiotics” (emmean: 1.55 vs 1.02, *p.adj* = 0.03) and “Naive” (emmean: 1.55 vs 1.04, *p.adj* = 0.03) groups (Figure 3B). Complementary analyses using Simpson diversity and Pielou’s evenness showed concordant patterns with Shannon diversity, while Observed Species Richness showed weaker associations, suggesting that the differences in α-diversity were primarily driven by changes in community evenness rather than species richness (Figure S9).

**Figure 3:**
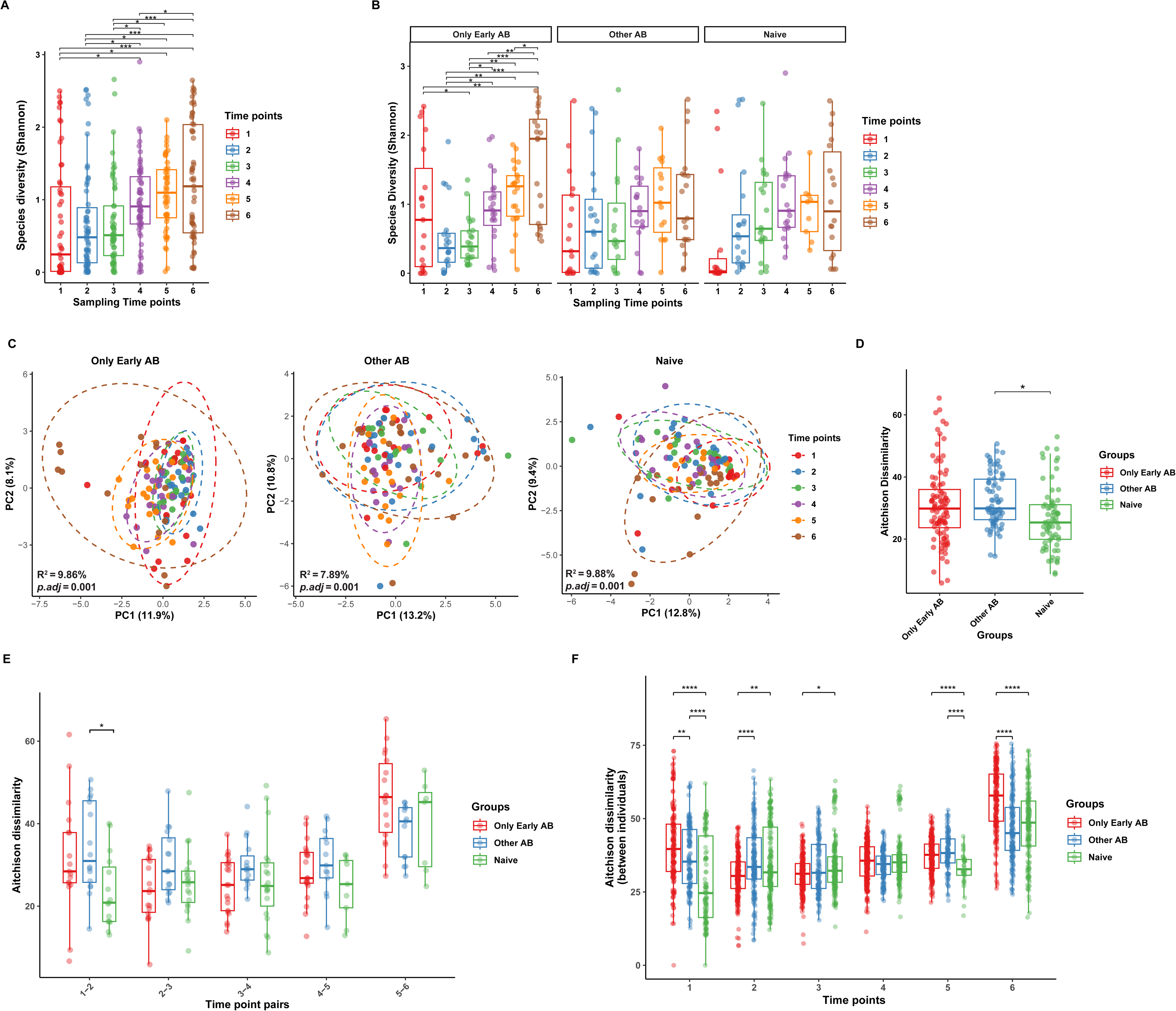
Effect of postnatal antibiotics on the nasopharyngeal microbial diversity, composition and stability. **(A, B)** Boxplots comparing species α-diversity (measured using the Shannon index) across sampling time points for all nasopharyngeal samples **(A)** and stratified by postnatal antibiotic exposure group **(B)**. Significant differences between time points were determined using an LMM approach that corrected for individual variation, followed by pairwise comparisons derived from estimated marginal means (emmeans) with BH correction. **(C)** Principal component analysis (PCA) plots based on Aitchison distances of CLR-transformed species abundances, stratified by postnatal antibiotic exposure group. Each point represents one sample, colored by time point, with ellipses representing 95% confidence intervals around the centroid of the microbiome composition for each group. Statistical significance, indicated by adjusted P-values (calculated using the Benjamini–Hochberg method) and effect sizes (R²), was assessed using a PERMANOVA and is displayed at the bottom left of each plot. **(D)** Boxplot comparing overall community stability, measured as within-infant Aitchison dissimilarity between consecutive sampling time points, across the three postnatal antibiotic exposure groups. **(E)** Boxplots comparing within-infant Aitchison dissimilarity stratified by consecutive time point pairs and postnatal antibiotic exposure group. **(F)** Boxplots comparing Aitchison dissimilarity between infants across sampling time points, stratified by postnatal antibiotic exposure group. In all boxplots, the central line represents the median, box indicates the IQR, and whiskers extend to 1.5× IQR. For panels **(D–F)**, pairwise Wilcoxon rank-sum tests with BH correction were used for between-group comparisons (\**p* < 0.05, \*\**p* < 0.01, \*\*\**p* < 0.001, \*\*\*\**p* < 0.0001). ns: not significant.

Next, we performed β-diversity analysis using permutational multivariate analysis of variance (PERMANOVA) to assess the impact of postnatal antibiotic exposure on the overall nasopharyngeal microbial community composition. After adjusting for gestational age (GA), no significant differences in community composition were observed between groups at any time point (PERMANOVA: all *p.adj* > 0.05; Supplementary Table E7; Figure S3A). Consistent with these community-level findings, differential abundance analysis identified no significantly differentially abundant taxa between groups at any time point, except a significant reduction in *Staphylococcus aureus* in the “Only Early Antibiotics” group compared to the “Naive” group at time point 2 (ANCOM-BC2: log fold change = −6.97, *p.adj* = 0.007; Supplementary Table E5), which did not remain significant after GA correction.

To examine longitudinal compositional dynamics within each group, we performed single-factor PERMANOVA with infant ID as strata to account for repeated measures. All three groups exhibited significant compositional change over time (all *p.adj* = 0.001; Figure 3C), consistent with the observed α-diversity trajectories. The “Only Early Antibiotics” group demonstrated significant compositional shifts from time point 2 onwards (time point 1 vs 2: R² = 3.6%, *p.adj* = 0.029), coinciding with ampicillin and gentamicin discontinuation. In contrast, the “Other Antibiotics” group showed no significant compositional change until time point 4 (time point 1 vs 4: R² = 5.2%, *p.adj* = 0.015), suggesting antibiotic-mediated suppression of early microbiome maturation. Compositional similarity between time points 4 and 5 across all groups (all *p.adj* > 0.05) suggests that the shared hospital environment may contribute to homogenization of the nasopharyngeal microbiome composition during the late NICU period, irrespective of antibiotic exposure.

We also investigated whether community stability and inter-individual compositional variation differed among antibiotic exposure groups over time. Community stability, measured by Aitchison dissimilarity between consecutive time points within each infant, was greatest in the “Naive” group, with significantly lower dissimilarity than the “Other Antibiotics” group (median: 25.3 vs 29.9; LMM: *p.adj* = 0.038; Figure 3D), driven primarily by greater instability at time points 1–2 (pairwise Wilcoxon: *p.adj* = 0.017; Figure 3E). Inter-individual compositional variation, quantified as Aitchison dissimilarity between all infant pairs within each group at each time point, was greatest in the “Only Early Antibiotics” group at birth (time point 1), significantly higher than both the “Other Antibiotics” (pairwise Wilcoxon: *p*.*adj* = 0.004) and “Naive” (*p.adj* < 0.001) groups (Figure 3F). In contrast, this group showed the lowest inter-individual variation at time point 2 (median: 39.6 vs 30.5; *p.adj* < 0.001 vs time point 1), becoming significantly more homogeneous than both the “Other Antibiotics” (*p.adj* < 0.001) and “Naive” (*p.adj* = 0.009) groups, suggesting that early antibiotic exposure drives microbial communities to become more similar across individuals, reflecting a homogenizing effect on nasopharyngeal microbiota development. All groups showed comparable inter-individual variation by time point 4 (all *p.adj* > 0.05; Figure 3F), consistent with the compositional convergence observed in β-diversity analysis. At six months corrected age, the “Only Early Antibiotics” group re-emerged as the most heterogeneous (median: 57.9), significantly exceeding both the “Other Antibiotics” (median: 45.1; *p.adj* < 0.001) and “Naive” (median: 48.6; *p.adj* < 0.001) groups.

When examining temporal dynamics within each group (Figure S4A, B), the “Only Early Antibiotics” group showed the most dynamic inter-individual variation, with no significant difference from the “Naive” group by time point 4 (all p*.adj* > 0.05), before re-emerging as the most heterogeneous at six months corrected age. The “Naive” group showed a significant increase in inter-individual variation from time point 1 to time point 4 (median: 24.6 vs 35.1; p*.adj* < 0.001), reflecting progressive community diversification in the absence of antibiotic exposure. The “Other Antibiotics” group maintained consistent inter-individual variation across time points 1–5 (all *p.adj* > 0.05), suggesting that ongoing antibiotic exposure suppressed natural community diversification. Across all groups, the greatest community instability was observed between time points 5–6, potentially driven by a combination of several environmental, infant-related, perinatal, and healthcare-associated factors.

### Factors influencing the nasopharyngeal microbiota development in preterm infants

To identify drivers of nasopharyngeal microbiota composition, we performed PERMANOVA analyses based on Aitchison distance and visualized the results using Principal Component Analysis (PCA) of centered log-ratio (CLR)-transformed data from samples collected from preterm birth to 6 months corrected age (∼7–10 months chronological age). Inter-individual variation had a pronounced effect on nasopharyngeal microbiota composition (R² = 33.84%, *p.adj* = 0.001; Figure S5A), highlighting the highly individualized nature of longitudinal microbiota changes and underscoring the importance of accounting for inter-individual variation in microbiome studies. Among clinical variables, postnatal age was the strongest significant predictor in single-factor PERMANOVA, whether described as postmenstrual age (PMA) (R² = 7.68%, *p.adj* = 0.002; Figure S5B), sampling time point (R² = 6.47%, *p.adj* = 0.002; Figure 4A) or days of life (DOL) (R² = 2.86%, *p.adj* = 0.002; Figure S5C). Antibiotic exposure group was also significantly associated with microbiota composition in single-factor analysis (R² = 1.28%, *p.adj* = 0.002; Supplementary Table E8).

**Figure 4.**
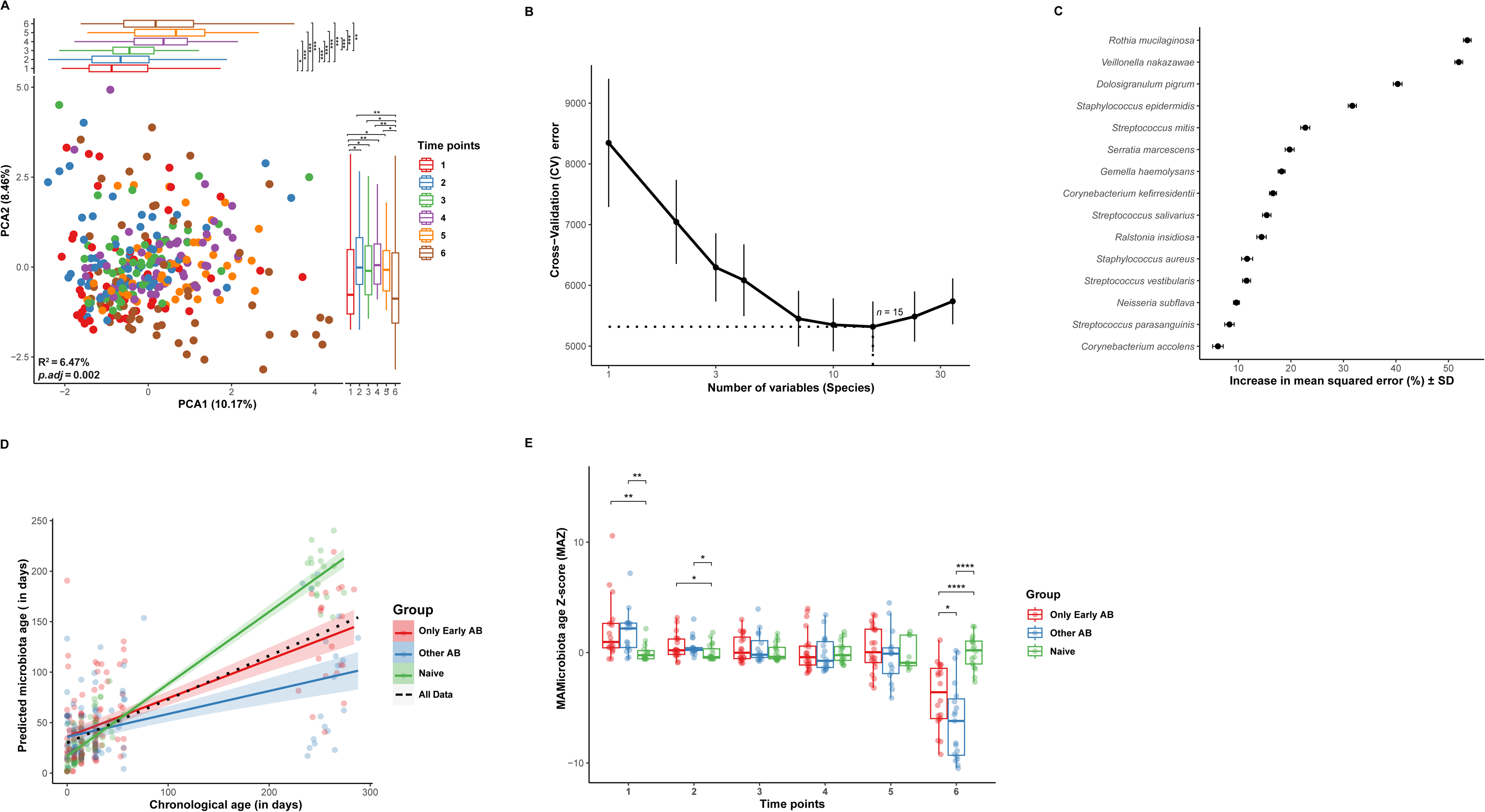
Nasopharyngeal microbiome composition of preterm infants evolves with age. **(A)** Principal Component Analysis (PCA) plot of the overall nasopharyngeal microbiota composition, based on CLR-transformed species abundances. Each point represents one sample (n = 331), colored by sampling time point. Boxplots on the bottom and right margins illustrate the distribution of samples along the first two principal components. Statistical significance was assessed using PERMANOVA (R² and BH-adjusted p-value displayed at the bottom left). Pairwise differences between time points in panel **(A)** were assessed using two-sided Wilcoxon rank-sum tests with BH correction (\**p* < 0.05; \*\**p* < 0.01; \*\*\**p* < 0.001; \*\*\*\**p* < 0.0001). **(B)** Fivefold cross-validation error as a function of the number of input species used to regress against infants’ chronological age in the “Naive” training set. The results indicate that 15 species are sufficient for random forest–based predictions of chronological age from the microbiota composition. Data are shown as the mean ± SD from 100 iterations. **(C)** The 15 most informative, age-discriminatory bacterial species identified by the random forest model, ranked in descending order by percentage increase in mean squared error (%IncMSE). Data are represented as the mean ± SD from 100 iterations. **(D)** Scatterplot showing the relationship between chronological age and the microbiome-predicted age (in days), with each dot representing one sample colored by postnatal antibiotic exposure group. Lines of best fit with 95% confidence intervals (shaded areas) were estimated by linear regression per group and for all data combined. **(E)** Microbiota-for-age Z-score (MAZ) at each sampling time point, stratified by postnatal antibiotic exposure group. In all boxplots, the center line denotes the median, box edges mark the 25th and 75th percentiles (IQR), and whiskers extend to 1.5 × IQR. Statistical significance is denoted as follows: \**p* < 0.05; \*\**p* < 0.01; \*\*\**p* < 0.001; \*\*\*\**p* < 0.0001.

Among baseline variables that differed between the three antibiotic exposure groups, gestational age (R² = 1.90%, *p.adj* = 0.002), birth weight group (R² = 1.17%, *p.adj* = 0.002) and antibiotic use during pregnancy (R² = 0.60%, *p.adj* = 0.009) were significantly associated with microbiota composition, while duration of ruptured membranes was not (R² = 0.31%, *p.adj* = 0.43; Supplementary Table E8). As birth weight group was collinear with gestational age and antibiotic use during pregnancy was partially reflected in the antibiotic exposure group classification, gestational age was selected as the only confounder and corrected for in subsequent PERMANOVA analyses.

In GA-corrected PERMANOVA, antibiotic exposure group (R² = 1.13%, *p.adj* = 0.001) and postnatal age remained significant. In PERMANOVA correcting for both gestational age and inter-individual variation (restricted permutations within each infant), only postnatal age remained significant (PMA: R² = 7.55%, *p.adj* = 0.003; sampling time point: R² = 6.52%, *p.adj* = 0.003; DOL: R² = 2.86%, *p.adj* = 0.003), while between-infant variables could not be assessed with this approach (all *p.adj* = 1.00; Supplementary Table E8). All clinical variables tested and their associations with microbiota composition across all PERMANOVA models are reported in Supplementary Table E8.

To further explore whether infant nasopharyngeal microbiota maturation is ubiquitously associated with age, we employed the Random Forest machine learning algorithm to regress microbial species relative abundances against the chronological age of preterm infants, as previously described ^36^. A total of 15 species were identified as the minimum number of variables required for accurate age prediction (Figure 4B). Using these 15 most informative age-discriminatory taxa, we trained the model on “Naive” infants to establish a reference maturation trajectory, refined, and validated the model to characterize nasopharyngeal microbiota development in preterm infants. The taxa most strongly associated with age in the nasopharyngeal microbiota were *Rothia mucilaginosa*, *Veillonella nakazawae* and *Dolosigranulum pigrum*, followed by *Staphylococcus epidermidis* and *Streptococcus mitis* (Figure 4C). This finalized sparse model was then applied to predict infant chronological age based on the relative abundances of these 15 taxa, producing a microbiota-derived predicted age that reflects microbiota maturity. A strong linear relationship was observed between microbiota age and chronological age of “Naive” preterm infants (Spearman: ρ = 0.965, *p* < 0.0001; Figure 4D), confirming the model’s accuracy in predicting preterm infant age. In contrast, the model showed substantially reduced predictive accuracy in antibiotic-exposed infants (“Only Early Antibiotics”: Spearman ρ = 0.696; “Other Antibiotics”: Spearman ρ = 0.506; both *p* < 0.001; Figure 4D), suggesting that microbiota maturation is delayed in these infants.

To further characterize when and to what extent antibiotic exposure delayed the expected microbiota maturation trajectory, we computed a microbiota-for-age Z-score (MAZ) for each sample as previously described ^36^, reflecting the deviation of predicted microbiota age from the antibiotic-naive reference trajectory at each time point. MAZ differed significantly between antibiotic exposure groups at early and late time points (Figure 4E). Surprisingly, at time points 1 and 2, both antibiotic-exposed groups exhibited significantly higher MAZ than the “Naive” group (“Only Early Antibiotics”: *p.adj* = 0.003 and 0.031; “Other Antibiotics”: *p.adj* = 0.003 and 0.024, respectively), potentially reflecting differences in early colonization patterns between groups relative to the antibiotic-naive reference. No significant between-group differences were observed at time points 3–5 (all *p.adj* > 0.05), consistent with the compositional convergence observed in β-diversity analyses. At six months corrected age, this pattern reversed as both antibiotic-exposed groups showed significantly lower MAZ than the “Naive” group (all *p.adj* < 0.001), with the “Other Antibiotics” group exhibiting the greatest deviation from the reference trajectory (*p.adj* = 3.12 × 10□□vs “Naive”), indicating delayed nasopharyngeal microbiota maturation in antibiotic-exposed infants at this developmental stage (Figure 4E).

### Developmental trajectory of the infant nasopharyngeal microbiota in early life based on community types

Using the Dirichlet multinomial mixture (DMM) method, we identified four optimal community type clusters (CT1–4) in the nasopharyngeal microbiota of all preterm infants (Figure S6A). These clusters accounted for 58.8% (CT1), 17.3% (CT2), 12.4% (CT3), and 11.5% (CT4) of all samples. These community types were mainly characterized by variations in the relative abundances of five bacterial genera: *Streptococcus*, *Serratia*, *Staphylococcus*, *Corynebacterium* and *Gemella* (Figure 5A, S6B). CT1 represented a mixed community state predominated by either *Streptococcus*, *Serratia* or *Staphylococcus*. CT2 was characterized by significantly higher *Staphylococcus* and *Corynebacterium* abundances compared to all other community types (pairwise Wilcoxon test: all *p.adj* < 0.05). CT3 exhibited significantly higher abundances of both *Streptococcus* and *Gemella* across all community types (all *p.adj* < 0.05). CT4 was distinguished by overwhelming *Serratia* dominance, significantly higher than all other community types (all *p.adj* < 0.001; Figure 5A, S6B).

**Figure 5.**
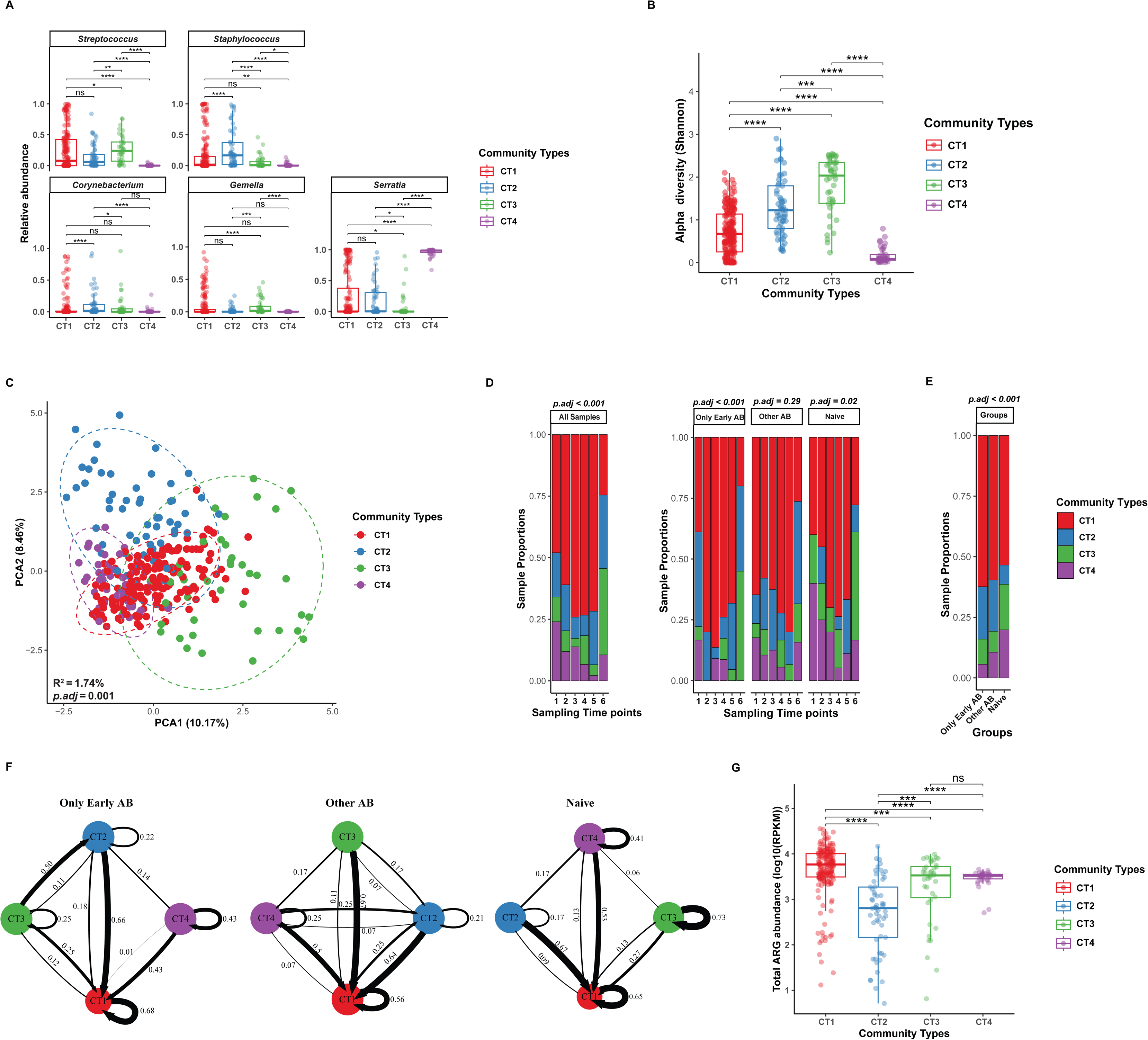
Identification of four community types in the nasopharyngeal microbiome of preterm infants. (**A)** Dirichlet multinomial mixture (DMM) analysis clustered the 331 samples into four distinct community types, visualized using Principal Component Analysis (PCA) based on Aitchison distances of species-level composition. Each point represents a single sample and is colored by community type. PERMANOVA R² and BH-adjusted p-value are displayed at the bottom left. (**B**) Box plots showing the relative abundance of the main contributing taxa at the genus level within each community type. (**C**) Boxplot of species diversity (Shannon index) in the nasopharyngeal microbiome samples of preterm infants, grouped by the four identified community types. (**D)** Stacked bar charts showing the proportions of community types at each sampling time point for all samples (left) and stratified by postnatal antibiotic exposure group (right) (n = 331). (**E)** Stacked bar chart showing the distribution of community types across the postnatal antibiotic exposure groups. BH-adjusted p-values from Pearson’s Chi-squared tests are displayed above panels **D** and **E**. (**F**) Stability and transition probabilities among the four microbial community types, evaluated *via* Markov chain modelling across different antibiotic exposure groups. Each node represents a community type identified by the DMM model, and arrow width is proportional to the transition probabilities among the community types. **(G)** Boxplots comparing the total antibiotic resistance gene (ARG) abundance (log10 RPKM) in nasopharyngeal samples from preterm infants, stratified by community types. In **B, C**, and **G**, the central line represents the median, box indicates the IQR, and whiskers extend to 1.5× IQR. Pairwise Wilcoxon rank-sum tests with BH correction were used to assess statistical significance. \**p* < 0.05, \*\**p* < 0.01, \*\*\**p* < 0.001 and \*\*\*\**p* < 0.0001. ns: not significant; CT: community type; RPKM: reads per kilobase per million.

We also found significant differences in microbial diversity (α-diversity, pairwise Wilcoxon test: all *p.adj* < 0.05; Figure 5B) and overall composition between all the identified community types, with β-diversity analysis confirming compositional separation (PERMANOVA: R² = 1.74%, *p.adj* = 0.001; Figure 5C). Sampling time point was significantly associated with community type clustering in the “Only Early Antibiotics” (Pearson’s Chi-squared: *p* < 0.001) and “Naive” (*p* = 0.026) groups (Figure 5D), but not in the “Other Antibiotics” group (*p* = 0.29), indicating an absence of structured temporal community state progression, consistent with the greater community instability observed in this group in earlier analyses. Additionally, community type distribution differed significantly between antibiotic exposure groups (Pearson’s Chi-squared: *p* < 0.001; Figure 5E), indicating that antibiotic exposure shapes the distribution of nasopharyngeal community states.

Next, we performed a Markov chain-based analysis to quantify the likelihood of transitions between community types in the early-life nasopharyngeal microbiota of preterm infants. The modeling revealed distinct transition patterns across postnatal antibiotic exposure groups (Figure 5F). CT1 (characterized by *Streptococcus*, *Serratia* and *Staphylococcus*) emerged as one of the most stable (*i.e.,* highest self-transition probability) states in the “Only Early Antibiotics” (self-transition: 0.68) and “Other Antibiotics” (0.56) groups, while CT3 (*Streptococcus*/*Gemella*-dominated) was the most stable state in the “Naive” group (self-transition: 0.73), suggesting that the health-associated commensal community state is maintained in the absence of antibiotic exposure. In the “Naive” group, CT4 (*Serratia*-dominated) transitioned predominantly toward CT1 (0.53), the mixed community state. In the “Only Early Antibiotics” group, CT3 transitioned predominantly toward CT2 (0.50) and CT4 showed equal probability of transitioning to CT1 or remaining in CT4 (0.43 each), both leading to lower diversity states. This is consistent with the transient decline in α-diversity and the disappearance of CT3 (the highest diversity community type), observed at time points 2 and 3 in this group (Figure 5D), underlining the impact of early antibiotic exposure on community state stability. On the other hand, CT3 in the “Other Antibiotics” group showed no self-transition (0.00 vs 0.73 and 0.25 in “Naive” and “Only Early Antibiotics”), transitioning entirely toward CT1 (0.67), indicating complete instability of the health-associated commensal state. This group also exhibited the lowest stability (self-transition probability) across most community types compared to both the “Naive” and “Only Early Antibiotics” groups, with more frequent transitions between states (Figure 5F), consistent with the greater community instability observed in this group in earlier analyses. Overall, these findings highlight the pronounced effects of postnatal antibiotic exposures on early-life microbial community dynamics, reducing overall community stability by making transient states less stable and promoting more frequent transitions across communities.

We further leveraged metagenomic sequencing data to investigate functional differences among community types by characterizing their antibiotic resistance (resistome) profiles ^27^. Striking differences were observed between the community types. CT1 and CT3 exhibited significantly higher total antibiotic resistance gene (ARG) abundance compared to CT2 (pairwise Wilcoxon test: all *p.adj* < 0.001; Figure 5G), suggesting that community states characterized by higher *Streptococcus* abundance harbor greater ARG loads in the infant nasopharyngeal microbiome.

### Prolonged signature of an outbreak on nasopharyngeal microbiota in preterm infants

A few months into our study, a clinical outbreak with *S. marcescens* was detected in the NICU. Six infants, including three from our study cohort, tested positive for *S. marcescens* in various clinical samples. Nasopharyngeal swab screenings were conducted on all hospitalized infants approximately two weeks after *S. marcescens* was first detected in our metagenomic results. The first screening yielded one positive case among study infants, while the second screening, conducted seven days later, found no positive nasopharyngeal cultures and the outbreak was declared over (Figure S7A). Environmental samples were negative, and no source of the outbreak was identified. However, metagenomic analysis of nasopharyngeal aspirate samples revealed persistent carriage of *S. marcescens* in infants hospitalized after the outbreak (Figure 6A). Notably, *S. marcescens*, which was detected early after birth (time point 1), persisted in over 90% (29/32) of preterm infants across all antibiotic exposure groups and remained detectable at ∼7–10 months of chronological age (time point 6), extending well beyond hospital discharge (Figure S7B). At time point 1, *S. marcescens* abundance was significantly higher in the “Naive” group compared to both the “Only Early Antibiotics” and “Other Antibiotics” groups (pairwise Wilcoxon test: both *p.adj* = 0.035), while no significant differences were observed at subsequent time points (all *p.adj* > 0.05; Figure 6B), likely reflecting the fact that all “Naive” infants were recruited following outbreak detection.

**Figure 6.**
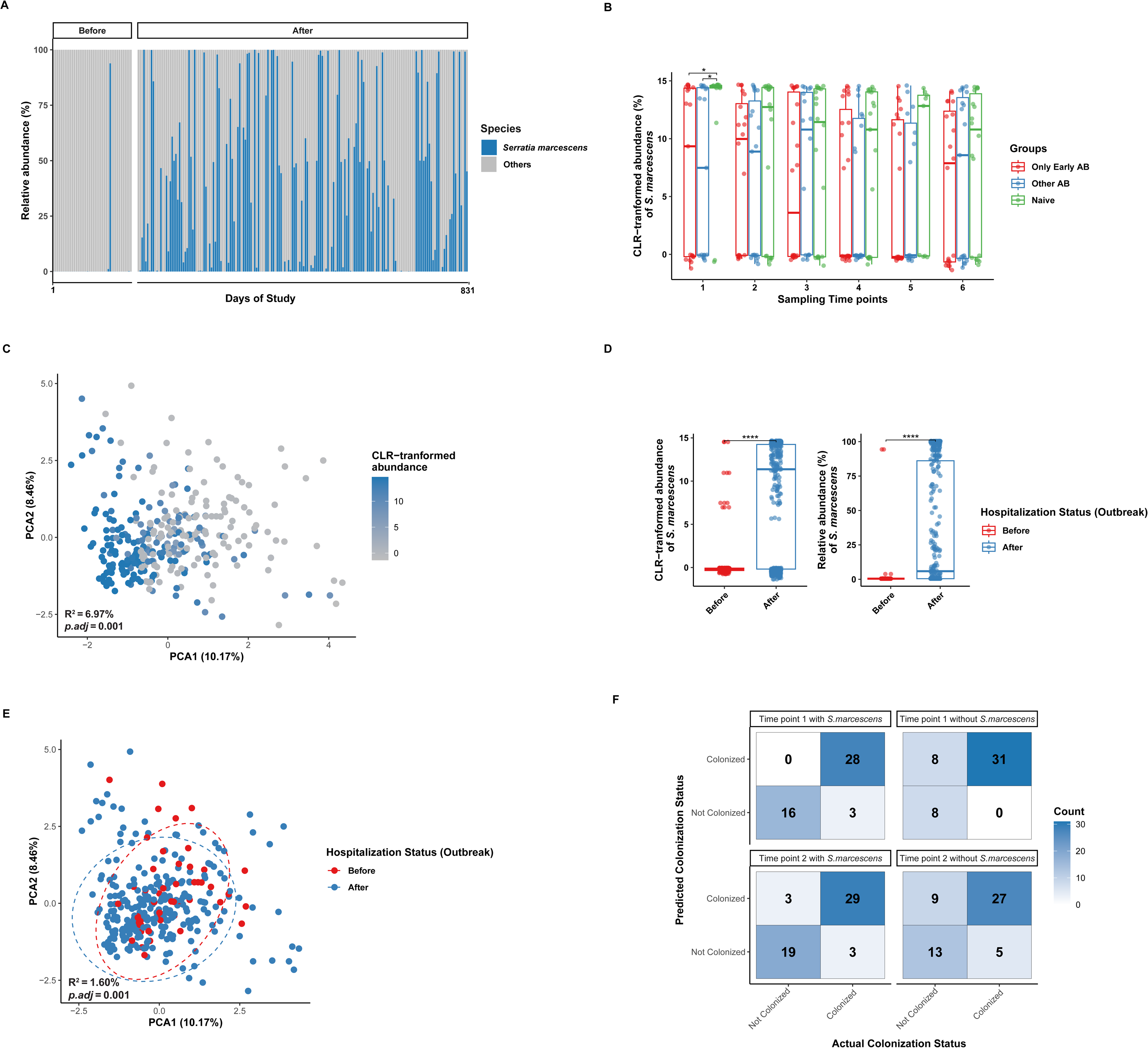
*S. marcescens* outbreak at the NICU. **(A)** Samples are shown in chronological order by sampling date. Each bar represents one sample, grouped by hospitalization status relative to the outbreak: “Before” (samples collected from infants discharged or transferred to a different NICU before the outbreak) and “After” (samples collected from infants hospitalized and discharged from the NICU after the outbreak). The relative abundance of *S. marcescens* is shown in blue, and the abundance of all other bacterial species is combined into the “Others” category (gray). **(B)** Boxplots illustrating the CLR-transformed abundance of *S. marcescens* across postnatal antibiotic exposure groups at each sampling time point (1–6). **(C)** PCA plot of the nasopharyngeal microbiota, colored according to the CLR-transformed abundance of *S. marcescens*. A continuous color gradient from gray (low) to blue (high) indicates CLR-transformed abundance, as shown in the legend. **(D)** Boxplots of the CLR-transformed (left) and relative abundance (%) (right) of *S. marcescens* in samples. Samples are grouped by hospitalization status (Before vs. After the outbreak). **(E)** PCA plot based on Aitchison distances of species-level abundances, colored by hospitalization status relative to the outbreak (Before vs After). Effect sizes (R²) and corresponding adjusted p-values (*p.adj*, BH-corrected) were calculated using PERMANOVA, as shown in the bottom left of the plot. Percentages on the axes indicate the variance explained by each principal component. **(F)** Random Forest confusion matrices for classifying preterm infants at six months corrected age as *S. marcescens* persistent or non-persistent, based on nasopharyngeal microbiota composition at time point 1 and time point 2, with and without *S. marcescens* included as a predictor. In **B** and **D**, the central line represents the median, box indicates the IQR, and whiskers extend to 1.5 × IQR. Pairwise Wilcoxon rank-sum tests with BH correction were used to assess statistical significance (\**p* < 0.05; \*\**p* < 0.01; \*\*\**p* < 0.001; \*\*\*\**p* < 0.0001).

Further, our analysis revealed that *S. marcescens* emerged as the dominant species, accounting for the most variation in overall microbiota composition (PERMANOVA: R² = 6.97%, *p.adj* = 0.001; Figure 6C). *S. marcescens* showed a significantly higher abundance in samples collected from infants hospitalized after the outbreak compared to those discharged or transferred to a different NICU prior to it (Wilcoxon rank-sum test: *p* < 0.001; Figure 6D). Prior to the outbreak being clinically recognized, *S. marcescens* was detected in only 7.3% of samples at low median relative abundance (1.76%). Following outbreak recognition, *S. marcescens* was detected in 65.9% of samples with a markedly higher median relative abundance (60.1%) among positive samples (Figure 6A), suggesting widespread acquisition together with expansion of low-level carriage that was already detectable before the outbreak was identified. Hospitalization status had a modest but significant influence on overall nasopharyngeal microbiota composition in single-factor PERMANOVA (R² = 1.75%, *p.adj* = 0.002), and remained significant after correcting for gestational age (R² = 1.60%, *p.adj* = 0.001; Figure 6E). Cross-sectional analysis per time point further revealed that compositional differences between hospitalization groups were already apparent at time points 1 and 2 (R² = 4.26% and 3.12%, *p.adj* = 0.039 and 0.048, respectively), and remained significant at six months corrected age (time point 6: R² = 3.75%, *p.adj* = 0.039; Figure S7C), suggesting that early nasopharyngeal microbiome composition reflects hospitalization status and may carry predictive information about long-term *S. marcescens* persistence.

To assess whether *S. marcescens* colonization confounded our primary findings, we performed sensitivity analyses correcting for *S. marcescens* CLR abundance. Antibiotic exposure group remained significantly associated with microbiota composition in β-diversity analysis (PERMANOVA: R² = 1.05%, *p* = 0.001), and the interaction between antibiotic exposure group and sampling time point in α-diversity remained significant (F = 1.987, *p* = 0.035), while *S. marcescens* itself was a highly significant independent predictor of both compositional variation (R² = 5.23%, *p* = 0.001) and Shannon diversity (F = 107.8, *p* < 2×10 ¹). CT4 was exclusively detected post-outbreak (38/38; 0 before outbreak) and was disproportionately prevalent in the “Naive” group (53% vs 9–14% in antibiotic-exposed groups). Excluding CT4, community type distribution among CT1–3 remained significantly different between antibiotic exposure groups (Fisher’s exact test: *p* = 0.013), confirming that antibiotic exposure independently influences microbiota composition beyond the effect of *S. marcescens* colonization, while also demonstrating the substantial and independent impact of the NICU outbreak on microbiota development in this cohort.

To understand the lasting effects of the outbreak on the nasopharyngeal microbiota, we sought to determine whether microbiome composition in early life could predict long-term *S. marcescens* persistence by identifying microbiome features at time points 1 and 2 that were predictive of *S. marcescens* colonization status at six months corrected age. Using a Random Forest classification model, we identified a sparse set of microbiome features that accurately predicted *S. marcescens* persistence (time point 1: 93.6%; time point 2: 88.5%; Figure 6F, S8A). To investigate whether microbiome community structure, independent of *S. marcescens* abundance, could predict persistence, models were retrained after excluding *S. marcescens* from the feature set, requiring only 10 microbiome features to retain substantial predictive accuracy (time point 1: 83.1%; time point 2: 73.7%; Figure S8A, B), indicating that early community composition independently informs colonization risk. Notably, models more accurately identified infants who would develop persistent nasopharyngeal *S. marcescens* colonization (sensitivity: 90.3–100%) than those who would remain uncolonized (specificity: 0–50%), suggesting that early nasopharyngeal microbiome composition is particularly informative of an infant’s susceptibility to *S. marcescens* persistence (Figure 6F). Collectively, these findings demonstrate that early nasopharyngeal microbiome composition independently predicts *S. marcescens* persistence and highlight the prolonged microbial signature established by *S. marcescens* in the nasopharynx of preterm infants, likely reflecting its acquisition during the NICU outbreak.

## DISCUSSION

The nasopharyngeal microbiota is a critical reservoir for microbes associated with respiratory tract infections, influencing immune development and disease susceptibility in infants ^10, 14, 17, 37^. This study provides a high-resolution longitudinal analysis of the preterm nasopharyngeal microbiota using whole metagenome sequencing. We demonstrated that the nasopharyngeal microbiota development from birth to 6 months corrected age (∼7-10 months chronological age) was characterized by dynamic and age-structured communities, with highly individualized patterns influenced by postnatal antibiotic exposure and hospitalization. Early-life antibiotic exposure (ampicillin + gentamicin) had pronounced ecological effects on microbial diversity, composition, stability, and community type transition dynamics, with some effects persisting to 6 months corrected age. A *Serratia marcescens* outbreak during the study provide a unique opportunity to assess its impact on microbiota development, revealing a persistent outbreak signature that, at ∼7–10 months chronological age, could be accurately predicted from nasopharyngeal microbiota composition in the first days of life.

After birth, preterm infants exhibited low bacterial abundance and diversity in the nasopharyngeal microbiota, followed by a sharp increase in the first few weeks and a more gradual rise for up to two months before stabilization. Similar trends have been described for term as well as preterm infants ^3, 7, 38, 39^, though our metagenomic approach offers enhanced taxonomic resolution at species level. Postnatal age was found to be associated with infant nasopharyngeal microbiome diversity, with a transient decline in α-diversity observed at time points 2 and 3 in the “Only Early Antibiotics” group, coinciding with ampicillin and gentamicin discontinuation. Infants in the “Other Antibiotics” group had significantly lower absolute bacterial abundance at two weeks after birth (time point 3) compared with both the “Only Early Antibiotics” and “Naive” groups. Overall, these findings suggest that postnatal antibiotic exposure transiently suppresses bacterial colonization and diversity in the nasopharyngeal microbiota of preterm infants.

Microbiome development in early life is a dynamic process, and transformations in the nasopharyngeal microbiota are expected, even in “Naive” infants. In our cohort, the nasopharyngeal microbiota of infants was predominantly colonized by five genera: *Serratia*, *Streptococcus Staphylococcus*, *Corynebacterium* and *Gemella*, forming different community types. At the species level, *S. marcescens*, *S. epidermidis*, *S. aureus*, and *S. mitis* were among the most abundant species identified. Carriage of specific bacterial species in the upper respiratory tract, such as methicillin-resistant *Staphylococcus aureus* (MRSA), has been closely linked to increased infection risks for newborns in the NICU ^40^. Among the 20 most abundant species, several were identified as World Health Organization (WHO) critical priority pathogens ^41^, including *Serratia marcescens*, *Klebsiella pneumoniae, Staphylococcus aureus,* and *Streptococcus pneumoniae*.

Because shotgun metagenomics enables higher taxonomic resolution than 16S rRNA studies, we were able to distinguish highly relevant pathogens from species generally considered commensals in immunocompetent individuals. For example, most *Streptococcus* species identified belonged to *Streptococcus mitis* across all ages. *S. mitis* is a close relative of *S. pneumoniae* and is considered a commensal that may provide protection against its pathogenic counterpart ^42^. In contrast, *S. pneumoniae*, a human colonizer with high pathogenic potential, appeared at lower abundances, reaching its highest proportion at 6 months corrected age, coinciding with when *S. pneumoniae* typically becomes more common in the nasopharynx of infants ^43^. Among the *Staphylococcus* species, we could distinctly identify *S. epidermidis*, initially more prevalent and peaking at time point 2, and *S. aureus*, a pathogen of greater concern, which became the dominant *Staphylococcus* from approximately four weeks (time point 4) of life onwards. Expanding our understanding of such species-level resolution of nasopharyngeal colonization dynamics may inform future surveillance and preventative strategies.

By six months corrected age, the nasopharyngeal microbiome shifted toward a more complex community structure, characterized by increased proportions of *Dolosigranulum pigrum* and *Moraxella catarrhalis*, alongside oral bacteria such as *Neisseria spp.* and *Veillonella spp*. *Dolosigranulum pigrum* was among the most informative age-discriminatory taxa identified by our machine learning model, consistent with previous studies ^3^. While *Corynebacterium* and *Dolosigranulum* are often associated with respiratory health benefits ^3, 6, 44^, their apparent lower abundance in the “Naive” group should be interpreted cautiously, as the disproportionate *S. marcescens* burden in this group, driven by the NICU outbreak, likely suppressed the relative abundance of other taxa through compositional effects rather than reflecting true biological differences. Early *Moraxella* colonization and low abundances of *Corynebacterium* and *Dolosigranulum* may predispose infants to chronic inflammation ^3, 45^, some studies suggest that *Moraxella*-dominant profiles are associated with community stability and fewer respiratory infections ^6^. Although our differential abundance analysis identified no significant differences in *Dolosigranulum* or *Moraxella* colonization between groups, the impact of antibiotic exposure on these health-associated taxa and their role in future respiratory health outcomes warrants investigation in larger cohorts with longer follow-up ^16, 46, 47^.

Inter-individual variability was the most significant factor influencing nasopharyngeal microbiome composition, consistent with previous findings in both term and preterm infants ^3, 15, 48^. In addition to individual differences, postnatal age also significantly impacted microbiome composition in our cohort, aligning with earlier studies ^3, 7, 49^. While cross-sectional analysis at birth did not reveal significant between-group compositional differences (R² = 4.33%, *p.adj* = 0.19), likely due to high inter-individual heterogeneity at this early stage, postnatal antibiotic exposure group remained independently associated with overall microbiota composition after GA correction (R² = 1.13%, *p.adj* = 0.001). The major community divergence between groups emerged during the first postnatal interval weeks, consistent with antibiotic exposure as the primary driver of subsequent microbiota differences. However, in contrast to Bosch et al. ^7^, we found no significant association between mode of delivery and nasopharyngeal microbiome composition in the first year of life. Similar findings have been published for the gut microbiome, with postnatal age having a greater influence than mode of delivery — both recognized as known factors shaping the microbiome composition of preterm infants ^36, 50^.

Early-life antibiotic exposure has been linked to several diseases ^16, 51, 52^. However, conflicting results have been reported in the literature regarding the impact of antibiotic exposure on nasopharyngeal microbiome composition in infants ^53^. In our cohort, early antibiotic treatment (ampicillin + gentamicin) had a pronounced homogenizing effect on the nasopharyngeal microbiome, characterized by reduced bacterial diversity, marked shifts in overall composition and microbiota stability, following antibiotic discontinuation. Moreover, Markov chain modeling revealed distinct community transition dynamics between groups. In the “Only Early Antibiotics” group, the health-associated *Streptococcus/Gemella*-dominated community type transitioned predominantly toward lower diversity states, while in the “Other Antibiotics” group, the health-associated community type lacked stability, transitioning entirely toward the mixed community state. Infants in the “Other Antibiotics” group, exposed to multiple classes of antibiotics (beta-lactam, aminoglycoside, glycopeptides, nitroimidazole, lincosamide), exhibited significantly lower temporal microbiome stability and more frequent transitions between community types compared to the “Naive” group, suggesting that exposure to multiple antibiotic classes may further disrupt the natural course of microbiota development typically observed in antibiotic-naive infants.

To further assess the impact of antibiotic exposure on microbiota maturation, we computed microbiota-for-age Z-scores (MAZ), which revealed an unexpected pattern of higher MAZ at time points 1 and 2 in antibiotic-exposed infants compared to the “Naive” group, potentially reflecting differences in early colonization patterns. By 6 months corrected age, this pattern reversed, with both antibiotic-exposed groups showing significantly lower MAZ than the “Naive” group, indicating delayed nasopharyngeal microbiota maturation at this developmental stage. This finding aligns with prior research on the gut microbiota suggesting that early-life antibiotic use disrupts microbial succession, with potential immune consequences ^54^. In the respiratory tract, changes in maturation of the nasopharyngeal microbiota have been correlated with increased risk of respiratory tract infections during the first year of life ^3^. Such relationships indicate that insights into the developmental dynamics of the nasopharyngeal microbiome could be key in identifying factors contributing to infection susceptibility and shaping strategies to reduce respiratory health risks in infancy. Beyond the microbiota differences observed here, even transient antibiotic-related alterations to the infant’s microbiome may have lasting consequences, including increased disease risk^17^, disrupted immune system development, and selection and spread of ARGs in both pathogenic and commensal bacteria ^36^. Implementation of stewardship actions focused on optimizing or reducing the initiation of early antibiotic therapy appears promising and has been adopted in some NICUs, though not yet for the smallest preterm infants ^55, 56^.

Beyond antibiotic exposure, the NICU environment itself represents a significant driver of upper respiratory tract microbiota and resistome development in preterm infants ^27, 57^, as evidenced in our cohort by the significant and persistent association between hospitalization status and nasopharyngeal microbiota composition. *S. marcescens*, an opportunistic pathogen prevalent in NICUs, can cause outbreaks and nosocomial infections in preterm infants ^58^. In a systematic review of published healthcare-associated outbreaks, *Serratia* species were identified as the third most frequent pathogen in neonatal units, accounting for 12.0% of documented NICU outbreaks and posing a significantly higher risk in neonatal settings compared to other intensive care units ^59^. Its persistence post-hospitalization and outbreak in our study underscores its role as a marker of hospital-acquired colonization, potentially influencing early-life microbiome development in preterm infants. It also coincided with a national outbreak of *S. marcescens* across 33 hospitals in Norway, without a common identifiable source ^60^. Although the outbreak was officially declared over after a short interval, our data indicate that *S. marcescens* persisted to the end of the study, highlighting the need for alternative long-term surveillance strategies to prevent silent persistence and re-emergence of outbreak strains in the future. Notably, previous studies do not report *Serratia* in the nasopharyngeal microbiota of term infants, and in preterm infants, it has been described only in very low abundance^38^. Random forest models demonstrated that early community structure alone, independently of *Serratia* abundance, predicted persistent *S. marcescens* colonization (TP1 accuracy: 83.1%), with as few as 10 microbiome features sufficient to retain substantial predictive accuracy. Markov transition analyses showed that the *Serratia*-dominated state (CT4) was self-reinforcing (self-transition: 0.25–0.43 across groups), and transitions out of CT4 led predominantly to the mixed community state (CT1) rather than the diverse health-associated CT3. Co-occurrence network analysis further showed that *S. marcescens* clustered exclusively with the opportunistic pathogens (*Acinetobacter pittii*, *Staphylococcus capitis*, and *Ralstonia insidiosa*) while health-associated commensals such as *Dolosigranulum pigrum* and *Corynebacterium* spp. occupied separate co-occurrence modules. Together, these findings suggest that persistent *S. marcescens* carriage occurs within a distinct microbial community context rather than in isolation, leaving a lasting imprint on community structure consistent with sustained ecological restructuring following NICU exposure. Additionally, resistome data from our previous study showed strong correlations between *S. marcescens* and intrinsic resistance genes conferring resistance to the empiric ampicillin-gentamicin regimen (*SST-1, AAC(6’)-Ic, tet(41)*), suggesting direct antibiotic selection pressure as a contributing mechanism. These observations support a dual model in which *S. marcescens* actively drives its own persistence through competitive exclusion and antibiotic resistance, while antibiotic-mediated depletion of competing taxa creates a permissive niche for its establishment and maintenance.

Our findings highlight two important clinical and ecological implications of the *S. marcescens* outbreak. First, nasopharyngeal carriage persisted well beyond the declared end of the outbreak, undetectable by routine clinical surveillance, demonstrating that outbreak strains can hide in plain sight within the microbiome, and underscoring the value of culture-independent metagenomic approaches for monitoring pathogen persistence in NICU settings. Second, early nasopharyngeal microbiome composition predicted *S. marcescens* persistence even after excluding *S. marcescens* from the prediction model, supporting the concept that persistent colonization is associated with the broader microbial context in which *S. marcescens* is established, although whether this reflects microbial interactions, shared ecological determinants, or intrinsic properties of *S. marcescens* remains to be determined.

A significant strength of our study is the serendipitous timing of the *S. marcescens* outbreak, which provided a unique chance to investigate the ecological impact of a nosocomial pathogen on the nasopharyngeal microbiota in a real-world setting. The study encompassed six sampling time points spanning from birth to approximately 7–10 months of chronological age in a homogeneous cohort of preterm infants born at 28–32 weeks GA at a single tertiary NICU. In addition, the inclusion of a “Naive” group is a particular strength, as many preterm infant studies lack an antibiotic-unexposed control group given that antibiotic administration is common practice in some hospitals for very and extremely low-birth-weight newborns ^61, 62, 63^. The consistent administration of early antibiotics, specifically ampicillin and gentamicin, ensured that comparisons are not diluted in significance due to variability in treatment, as different antibiotics often have distinct impacts on the microbiota ^64^. Moreover, we followed recommended low-biomass research practices for our nasopharyngeal samples ^65^, including sequencing negative and positive controls and rigorous *in silico* contaminant removal to minimize contamination and bias, ensuring reliable and accurate findings. Infants remained in a controlled NICU environment for most sampling periods, had no daycare exposure before the final sampling, and followed a consistent vaccination schedule, minimizing external influences ^14, 17^. Additionally, we used nasopharyngeal aspirates instead of nasal swabs to reduce contamination from the anterior nostril microbiome ^66^. To our knowledge, this is one of the first studies to longitudinally track the development of the preterm infant nasopharyngeal microbiome with species-level metagenomic resolution, offering detailed insights into microbial dynamics.

Our study has some limitations. Firstly, we focused on known factors influencing the microbiome and may have overlooked other relevant variables. Notably, we did not assess potential microbial sources such as hospital surfaces, healthcare personnel, or maternal microbiota, which could contribute to infant nasopharyngeal microbiome development. Additionally, the study did not investigate some lifestyle factors known to influence early microbiome development, such as probiotics, respiratory infections, sibling or pet exposure, and vaccination history. Furthermore, our findings do not by themselves establish a causal relationship between antibiotic treatment and the observed ecological effects. Further mechanistic studies, including animal, ex vivo, and in vitro models, may help clarify the underlying mechanisms. However, our findings strongly suggest a direct biological impact of antibiotics, as the microbiota composition in the treatment group showed significant alterations following exposure, with some effects persisting to six months corrected age. While gestational age differed significantly between antibiotic exposure groups and was accounted for in compositional analyses where applicable, postnatal antibiotic exposure group remained independently associated with microbiota composition after this correction, indicating that the observed microbiota differences between groups are not fully explained by differences in gestational age alone. Nevertheless, maturational and clinical differences between antibiotic-exposed and “Naive” infants likely extend beyond what can be fully captured by statistical adjustment, and should be considered when interpreting between-group differences in nasopharyngeal microbiota development. Additionally, the temporal overlap between the *S. marcescens* outbreak and the recruitment of most “Naive” infants may have contributed to their distinct microbiota profiles, and these factors cannot be fully disentangled. The absence of a term-born control group limits direct comparison of nasopharyngeal microbiota development between preterm and term infants. However, this was not feasible given the separate care settings and markedly shorter hospital stays of term-born infants. The study’s single-center design may also limit the broader applicability of our findings. The study was conducted during the COVID-19 pandemic, a period associated with substantial changes in social contact patterns and microbial exposures in infants. Although NICU infection-control practices remained largely unchanged during this period, we cannot exclude the possibility that pandemic-related societal measures influenced microbiota development after hospital discharge and therefore contributed to some of the observed longitudinal patterns. Strain-level diversification of *S. marcescens* and other key taxa was not assessed in the current study and represents an important direction for future investigation. Additionally, concurrent processing of each infant’s samples (all samples in the same batch) may have reinforced individual microbiome signatures. Despite equimolar library pooling prior to WMS, variation in raw and bacterial reads was observed. Nonetheless, we rigorously accounted for inter-individual variation and employed analytical methods that mitigate sensitivity to sequencing depth while addressing the compositional nature of microbiome data ^67, 68^, ensuring the reliability and robustness of our findings.

Overall, this study highlights the significant influences of age, outbreak events, and postnatal antibiotic exposure on the developmental dynamics of the nasopharyngeal microbiota in preterm infants. The pronounced alterations in microbial diversity, composition, stability and maturation observed in response to antibiotic exposure, along with the enduring impact of microbial signatures from outbreak events, underscore the importance of understanding these factors in shaping the microbiome in early life. Preterm infants are exposed to multiple factors known to shape the microbiota across various body niches ^69, 36, 70, 71^. There is also growing evidence that respiratory health outcomes may be influenced by the intestinal and oral microbiota ^9, 15, 72^. Future studies are warranted to investigate the interconnected colonization dynamics of these body niches during infancy. Additionally, ongoing efforts to develop strategies for restoring microbial balance and minimizing unnecessary antibiotic use during this critical phase of development represent a valuable pathway for preventing disturbances in microbiota-host interactions ^73, 74^, which may have potential implications for health.

## Ethical statement

The study was performed in accordance with the Declaration of Helsinki and approved by the Hospital’s Data Protection Officer and the Regional Committee for Medical and Health Research Ethics-South East, Norway (2018/1381 REKD).

## Consent statement

Written informed consent was obtained from the infant’s parents. The participants received no compensation.

## Data availability

The metagenomic sequencing data (clean reads *i.e.,* after quality filtering and removal of human DNA) generated from all nasopharyngeal samples are publicly available on the NCBI Sequence Read Archive database under BioProject ID: <u>PRJNA1009231</u>.

## Acknowledgments

We would like to thank the parents of participating infants, and the clinical staff at the Neonatal Intensive Care units at Ullevål hospital for their assistance with sample collection. The sequencing service for this work was provided by the Norwegian Sequencing Centre (www.sequencing.uio.no) and the computations were performed on resources provided by Sigma2 -the National Infrastructure for High Performance Computing and Data Storage in Norway.

## Competing interests

The authors declare that the research was conducted in the absence of any commercial or financial relationships that could be construed as a potential conflict of interest.

## Author contributions

FCP and KH conceived the study. All authors contributed to the design of the study and interpretation of data. PR and KH included the infants, collected samples and patient metadata. PR, GS and HAA processed the samples under FCP supervision. AD performed all whole-metagenome sequencing data processing, bioinformatic analyses, statistical analyses, and data visualization. PR performed clinical and demographic data curation and statistical analyses. PR and AD wrote the manuscript. FCP provided overall scientific supervision of the study. All authors discussed the results, critically revised the manuscript, and agreed to the published version of the manuscript.

## Funding

This work was primarily funded by the Research Council of Norway (RCN) project numbers 273833 and 322375, and by the Olav Thon Foundation, with additional support from the Faculty of Dentistry at the University of Oslo and by Oslo University Hospital.

---

**Figure S1**. Overview of obtained samples and sampling time points in the study cohort. **(A)** Nasopharyngeal aspirate (NPA) samples collected from each infant (left y-axis) are connected by horizontal lines. Infants are organized by postnatal antibiotic exposure group (right y-axis). The x-axis represents the day of life (DOL). In total, 369 NPA samples were obtained from 66 infants within the first six months of corrected age. **(B)** NPA samples were collected at six designated sampling time points (y-axis). If a sample could not be obtained at the designated time, a new sample was collected as soon as possible, leading to greater variation in DOL for each time point. Variation in actual collection dates additionally reflects timing of antibiotic initiation and discontinuation. Horizontal bars indicate the DOL range for each sampling time point and the vertical line within each bar denotes the mean. Mean (SD) values are listed on the right. **(C)** Scatter plot showing the correlation between bacterial abundance measured by qPCR (log 16S qPCR, ng) and estimated bacterial read counts (in millions) from MetaPhlAn4-based taxonomic profiling. Each dot represents a single sample. ρ = Spearman’s correlation coefficient.

**Figure S2.** Stacked bar plots showing the mean relative abundances of bacterial taxa across sampling time points and postnatal antibiotic exposure groups. **(A–B)** Mean relative abundances of the five most abundant phyla by sampling time point **(A)** and stratified by postnatal antibiotic exposure group **(B)**. **(C–D)** Mean relative abundances of the 15 most abundant genera by sampling time point **(C)** and stratified by postnatal antibiotic exposure group **(D)**. All remaining phyla and genera not among the most abundant are grouped into the “Others” category.

**Figure S3.** PCA plots based on Aitchison distances of CLR-transformed species abundances, visualizing the overall nasopharyngeal microbial community composition of preterm infants at each sampling time point **(A–F)**. Each point represents a sample and is colored by postnatal antibiotic exposure group. Ellipses represent the 95% confidence interval for each group. Effect sizes (R²) from permutational multivariate analysis of variance (PERMANOVA), along with corresponding adjusted P-values (*p.adj*, calculated using the Benjamini–Hochberg method), are shown at the bottom right of each plot.

**Figure S4.** Boxplots illustrating the temporal stability of the nasopharyngeal microbiota, stratified by postnatal antibiotic exposure group. **(A)** Boxplots comparing within-infant Aitchison dissimilarity between consecutive time point pairs (1–2, 2–3, 3–4, 4–5, 5–6) across postnatal antibiotic exposure groups. **(B)** Boxplots comparing Aitchison dissimilarity between infants at each time point, stratified by postnatal antibiotic exposure group. In all boxplots, the central line represents the median, box indicates the IQR, and whiskers extend to 1.5 × IQR. Pairwise Wilcoxon rank-sum tests with BH correction were used (\**p* < 0.05; \*\**p* < 0.01; \*\*\**p* < 0.001; \*\*\*\**p* < 0.0001).

**Figure S5.** Individual variability and postnatal age significantly influence the nasopharyngeal microbiome composition in preterm infants. PCA plots (based on CLR-transformed species abundances) illustrate β-diversity of the nasopharyngeal microbiota. Panels **(A)**, **(B)**, and **(C)** show the effects of individual variability (infant ID), postmenstrual age (PMA), and days of life (DOL), respectively. Each point represents one sample (n = 331), colored by infant ID **(A)**, PMA **(B)**, or DOL **(C)**. Effect sizes (R²) and corresponding adjusted P-values (*p.adj*, Benjamini–Hochberg correction) were calculated using PERMANOVA and are displayed at the bottom left of each panel.

**Figure S6.** Dirichlet multinomial mixture (DMM) analysis of nasopharyngeal samples reveals four distinct community types. **(A)** Model fit statistics (AIC, BIC, and Laplace approximation) across DMM components (k = 1–9), indicating four as the optimal number of community types. **(B)** Top 10 driver genera for each DMM-based community type, ranked by their contribution to each community type (fitted genus weight).

**Figure S7. (A)** Overview of the 13 NICU infants (y-axis) screened at least once during the two main clinical screening days (x-axis). Despite negative clinical screening results, metagenomic profiling identified *S. marcescens* reads in nine infants, with relative abundances ranging from <1.3% to 85.0%. **(B)** Timeline of *S. marcescens* detection in nasopharyngeal samples from each infant across sampling time points, where each point represents a sample, colored by whether it was collected before or after the outbreak, and point shapes indicate whether *S. marcescens* was identified in metagenomic data. **(C)** Principal Components Analysis (PCA) plots of β-diversity (Aitchison distances) at each time point (1–6), with each point representing a sample and colored by hospitalization status relative to the outbreak. Adjusted p-values (Benjamini–Hochberg correction) and effect sizes (R²) for all time points were calculated using PERMANOVA and summarized in the table on the right.

**Figure S8.** Random Forest model performance for predicting *S. marcescens* colonization at six months corrected age. **(A)** Cross-validated accuracy of full and sparse Random Forest models at time point 1 and time point 2, with and without *S. marcescens* included as a predictor. **(B)** Top 10 species contributing most to classification at time point 1 and time point 2, with and without *S. marcescens*, ranked by variable importance.

**Figure S9.** Complementary α-diversity metrics stratified by postnatal antibiotic exposure group. **(A)** Spearman correlation between bacterial read counts and Shannon diversity across all samples (ρ = −0.215; *p* < 0.001). **(B–D)** Boxplots comparing Simpson diversity (B), Pielou’s evenness **(C)**, and Observed Species Richness **(D)** across sampling time points, stratified by postnatal antibiotic exposure group. In all boxplots, the central line represents the median, box indicates the IQR, and whiskers extend to 1.5 × IQR. Pairwise comparisons were derived from estimated marginal means (emmeans) of the fitted LMM with BH correction (\**p* < 0.05; \*\**p* < 0.01; \*\*\**p* < 0.001; \*\*\*\**p* < 0.0001).

**Supplementary Table E1.** Metadata for 331 nasopharyngeal samples from 66 preterm infants included in microbiome analyses, including clinical variables and sample-level information.

**Supplementary Table E2.** Overview of sample processing. Summary of nasopharyngeal aspirate samples collected from preterm infants across different stages of the study, including sampling, DNA extraction, sequencing, and microbiome analysis. The table shows the number and proportion of samples excluded at each step for all infants (n = 66), antibiotic-naive infants (n = 21), and antibiotic-exposed infants (n = 45) across six time points. DOL: days of life.

**Supplementary Table E3.** DNA quantification, library preparation, and sequencing statistics for each sample. The table summarizes Qubit and Femto DNA measurements, sequencing read counts (total, trimmed, human, and clean reads), and estimated bacterial and eukaryotic reads derived from MetaPhlAn4-based taxonomic profiling.

**Supplementary Table E4.** Decontamination analysis results per species, showing frequency and prevalence of contaminant and non-contaminant bacteria identified during glycerol blank and DNA extraction (PMA) controls using the decontam package. freq: frequency; prev: prevalence; p.freq: p-value frequency method; p.prev: p-value prevalence method; p: combined p-value.

**Supplementary Table E5.** ANCOM-BC2 differential abundance analysis results. Log fold changes (LFC) and BH-adjusted *p*-values (*p.adj*) for pairwise comparisons of each antibiotic exposure group against the “Naive” reference group, computed using ANCOM-BC2 without and with correction for gestational age (GA). Results are organized by sampling time point (time points 1–6, one sheet per time point). Within each sheet, columns highlighted in blue correspond to results without GA correction and columns highlighted in green correspond to results with GA correction. Significant results (*p.adj* < 0.05) are highlighted in yellow and listed at the top of each sheet. LFC: log fold change; GA: gestational age.

**Supplementary Table E6.** Likelihood ratio test (LRT) results assessing associations between clinical variables and Shannon α-diversity and absolute bacterial abundance (log10 bacterial DNA by RT-PCR), using linear mixed-effects models with infant identity as a random effect. LRT p-values and significance levels are reported for each variable. ns: not significant.

**Supplementary Table E7.** Pairwise PERMANOVA results comparing nasopharyngeal microbiota composition between antibiotic exposure groups at each sampling time point (1– 6), adjusted for gestational age, along with within-group PERMANOVA assessing overall and pairwise compositional change over time, with infant ID as strata to account for repeated measures. All analyses based on Aitchison distances (999 permutations) with BH correction. Significance: \**p* < 0.05; \*\**p* < 0.01; \*\*\**p* < 0.001; ns: not significant. R²: variance explained; F: F-statistic; AB: antibiotic; TP: time point; GA: gestational age.

**Supplementary Table E8.** Single-factor and multifactorial PERMANOVA results (R², F-value, p-value, and BH-adjusted p-value) examining associations between clinical variables and overall nasopharyngeal microbiota composition across all time points, based on Aitchison distances (999 permutations). Three analytical approaches were applied: single-factor PERMANOVA; GA-corrected PERMANOVA adjusting for gestational age group (by = ‘margin’); and within-infant restricted PERMANOVA (strata = infant ID, by = ‘margin’). Note that between-infant constant variables yield *p* = 1.00 in the restricted permutation model by statistical necessity. Aitchison distances were used with 999 permutations and *p*-values were corrected using the Benjamini–Hochberg method. Significance: *** *p* ≤ 0.001; ** *p* ≤ 0.01; * *p* ≤ 0.05. MOB: mode of birth; BW: birth weight; PMA: postmenstrual age; DOL: days of life; IAP: intrapartum antibiotic prophylaxis; ROM: rupture of membranes; AB: antibiotics; LOS: late-onset sepsis.

**Supplementary Table E9.** Collinearity assessment between clinical variables. Categorical-categorical associations were evaluated using Cramér’s V (Chi-square or Fisher’s exact test); continuous-categorical associations using Kruskal-Wallis tests; and continuous-continuous associations using Spearman correlation. P-values adjusted using Benjamini–Hochberg correction. Abbreviations as in Table E4.

## Notes

### Competing Interest Statement

The authors have declared no competing interest.

### Author Declarations

The study was performed in accordance with the Declaration of Helsinki and approved by the Hospitals Data Protection Officer and the Regional Committee for Medical and Health Research Ethics-South East, Norway (2018/1381 REKD).

### Summary of Updates

We have revised our manuscript and re-analyzed the data. Most notably, we reprocessed the entire metagenomic dataset using MetaPhlAn4 with a more stringent taxonomic assignment threshold. MetaPhlAn4 was specifically developed to improve species-level taxonomic resolution through a substantially revised marker database with greater marker uniqueness and fewer spurious cross-species assignments. We also adopted a more stringent (default) abundance estimation threshold (--stat_q = 0.2 instead of 0.05). The lower threshold had originally been selected to maximize sensitivity for detecting low-abundance spike-in species included in our validation experiments, whereas the more stringent threshold reduces low-confidence assignments among closely related taxa. This updated analysis resulted in a more conservative species profile. Serratia nematodiphila and Enhydrobacter aerosaccus were no longer detected in any sample, and the number of Streptococcus species identified was reduced, indicating that these previous assignments most likely reflected low-confidence classifications.

