## Supplementary Methods for "Outbreak and Postnatal Antibiotic Exposures Drive the Development Trajectory of the Nasopharyngeal Microbiota in the First Year of Life"

### MATERIALS & METHODS

**Metagenomic DNA extraction**

Frozen nasopharyngeal aspirate samples were thawed on ice. The starting volume for all samples was approximately 2 ml. Host cells were chemically lysed and extracellular DNA removed by enzymatic degradation (MolYsis™ Basic5, Molzym). Samples were then spiked with 20 µl of Spike-in Control II Low Microbial Load (Catalog D6321 & D6321-10, ZymoBIOMICS™, Irvine, CA), immediately followed by DNA extraction (MasterPure™ Gram Positive DNA Purification Kit, Epicentre) ^1^. Extracted DNA was eluted in 35 µl, quantified using the Qubit™ dsDNA HS kit on a Qubit 4.0 Fluorometer (Invitrogen, Thermo Fisher Scientific, Waltham, MA) and stored at −80°C for up to four weeks before library preparation.

**Bacterial and Host DNA quantification**

Human and microbial DNA fractions were quantified using Zymo Femto™ Quantification kits (Zymo E2005 and E2006, ZymoBIOMICS™) by real-time qPCR using 1 µl sample volume per manufacturer's instructions. From the 16S rRNA qPCR results, the average yield of bacterial DNA from the spike-in mock was deducted from bacterial DNA yields of patient samples to determine each sample's own bacterial load. Values were log-transformed prior to analysis.

**Controls for low-biomass samples**

Six aliquots of 20 µl Spike-in Control II (ZymoBIOMICS™) were extracted with MasterPure™ Gram Positive DNA Purification Kit (Epicentre) in two independent experiments and used as positive controls (expected yield 0.4 ng). Glycerol saline solution samples from each cryopreservation batch (n = 5) and reagent blanks (n = 5) were extracted as negative controls. DNA yield from negative controls was extremely low or undetected (n = 10; mean = 0.00043 ng/µl) as measured by real-time qPCR.

**Metagenomic library preparation**

Nextera DNA Flex kit (Illumina Inc., San Diego, CA) was used for library preparation following manufacturer’s instructions. Input yield was set to 10 ng DNA according to Qubit results, and all available extracted DNA was used in samples that yielded less than 10 ng according to both Qubit and PCR measurements. Twenty-four samples from 16 infants were excluded from library preparation due to negative 16S rRNA qPCR values, determined by subtracting their bacterial DNA yield from the average yield of positive spike-in controls (Supplementary Table E2). Twelve PCR amplification cycles were used for metagenomic library preparation. This cycle number was selected based on our previous optimization study and in accordance with the manufacturer's preparation protocol for the DNA input used in this study ^1^. Library concentration and purity were measured using Qubit™ dsDNA HS kit (Invitrogen) and Bioanalyzer 2100 (Agilent, Santa Clara, CA). Libraries of the remaining 345 nasopharyngeal samples proceeded to WMS in one pool with 5 nM concentration. Libraries of positive (n = 6) and negative controls (n = 10) were pooled separately at 9.4 nM concentration.

**Resistome profiling**

Quality-filtered non-host reads were mapped to the Comprehensive Antibiotic Resistance Database (CARD, v.3.2.2) ^2^ using Bowtie2 ^3^ with the *--very-sensitive-local* parameter. Mapped reads were sorted, indexed and quantified using SAMtools ^4^ and BEDTools ^5^. ARGs with ≥80% sequence coverage were considered detected. Read counts were normalized for gene length and bacterial sequence abundance by calculating reads per kilobase per million bacterial reads (RPKM) for each ARG.

***In silico* identification and removal of contaminants**

Potential contaminants were identified using both the prevalence and frequency-based methods of the decontam R package (v.1.16.0) ^6^ with the default threshold (p-score = 0.1), applied independently within each batch of glycerol blank and DNA extraction controls. Identified contaminants were removed from species-level abundance tables prior to all downstream analyses.

**Covariate selection**

To identify potential confounders, we assessed collinearity between all clinical variables using Cramér's V for categorical-categorical associations, Kruskal–Wallis tests for continuous-categorical associations, and Spearman correlation for continuous-continuous associations (Supplementary Table E9). Variables were considered confounders only if they both (1) differed significantly between antibiotic groups at baseline and (2) were significantly associated with nasopharyngeal microbiota composition in single-factor PERMANOVA models.

At baseline, antibiotic-exposed infants differed significantly from the “Naive” group in gestational age, birth weight, antibiotic use during pregnancy, and duration of membrane rupture (Table 1). Of these, gestational age group (R² = 1.90%, *p.adj* = 0.002), birth weight group (R² = 1.17%, *p.adj* = 0.002) and antibiotic use during pregnancy (R² = 0.60%, *p.adj* = 0.009) were significantly associated with microbiota composition in single-factor PERMANOVA, while duration of membrane rupture was not (*p.adj* = 0.43; Supplementary Table E8). Mode of birth, sex, and ROM duration were not significantly associated with microbiota composition (all *p.adj* > 0.05; Supplementary Table E8) and were therefore not considered as covariates.

Gestational age group was the only variable meeting both criteria without collinearity with the main predictor (antibiotic group) and was therefore selected as the sole covariate. Birth weight group was excluded due to high collinearity with gestational age group (Cramér's V = 0.44; *p.adj* = 0.004). Prenatal antibiotic exposure, although differing at baseline and associated with microbiota composition, was excluded due to moderate collinearity with antibiotic group assignment (Cramér's V = 0.54; *p.adj* = 0.004), which would risk underestimating the true effect of postnatal antibiotic exposure. Gestational age was consequently corrected for in the majority of downstream analyses where applicable, including between-group beta diversity PERMANOVA (Supplementary Table E7), differential abundance analysis (ANCOM-BC2; Supplementary Table E5), and sensitivity analyses correcting for *S. marcescens* CLR abundance. It was not included as a covariate in alpha diversity analyses, as gestational age was not significantly associated with Shannon diversity in likelihood ratio tests (Supplementary Table E6).

**Alpha diversity analysis**

Alpha and beta diversity metrics were calculated using the phyloseq R package (v.1.40.0) ^7^. Alpha diversity was assessed using the Shannon diversity index as the primary metric, integrating both species richness and evenness. Complementary metrics including Simpson diversity, Pielou's evenness, and observed species richness were additionally calculated. Association of clinical variables with Shannon diversity was screened using likelihood ratio tests (LRT) comparing full and null linear mixed-effects models (LMMs) with infant identity as a random effect, implemented in the lme4 package (v.1.1.35.1) ^8^. Shannon diversity was subsequently modelled using an LMM with time point, antibiotic exposure group, and their interaction as fixed effects and infant identity as a random effect. Pairwise comparisons across time points within each antibiotic exposure group, and between groups at each time point, were derived from estimated marginal means (emmeans) of the fitted model with BH correction, implemented in the emmeans package (v.1.10.5) ^9^. To assess potential sequencing depth bias, Spearman correlation between bacterial read counts and α-diversity indices revealed that Shannon (ρ = −0.215, p < 0.001) and Simpson (ρ = −0.229, p < 0.001) diversity showed only weak associations with library size, while Observed Species Richness (ρ = +0.268, p < 0.001) and Pielou's evenness (ρ = −0.517, p < 0.001) showed greater sensitivity to sequencing depth, supporting the selection of Shannon as the primary diversity metric. The time point × antibiotic exposure group interaction was additionally tested for Simpson diversity (F = 1.916, p = 0.043), Pielou's evenness (F = 2.066, p = 0.028), and Observed Species Richness (F = 1.345, p = 0.207), confirming that the reported α-diversity patterns were robust across different diversity metrics and primarily driven by changes in community evenness rather than species richness

**Beta diversity and PERMANOVA analysis**

Microbiota compositional differences were calculated using Aitchison distance on centered log-ratio (CLR)-transformed species-level abundance data. Compositional differences were visualized using principal component analysis (PCA) of CLR-transformed data. Associations between clinical variables and microbiota composition were assessed using PERMANOVA (adonis2 function, vegan v.2.6.4 ^10^, 999 permutations) in three sequential steps. First, each clinical variable was tested independently against microbiota composition. Second, variables significant in step 1 were retested after correcting for gestational age (by = 'margin') to identify variables with independent associations beyond gestational age effects. Third, variables were retested after additionally restricting permutations within each infant (strata = infant ID, by = “margin”) to account for repeated measures and inter-individual variation. Note that between-infant constant variables (antibiotic exposure group, gestational age, outbreak status) yield *p* = 1.00 in step 3 by statistical necessity, as restricted permutation designs cannot generate a valid null distribution for variables that do not vary within infants. P-values were BH-adjusted across all variables at each step. Cross-sectional PERMANOVA was additionally performed at each time point separately to assess compositional differences between antibiotic exposure groups, correcting for gestational age (by = “margin”), with p-values BH-adjusted across time points.

**Microbial co-occurrence network analysis**

Microbial co-occurrence networks were inferred using SparCC correlations implemented in the SpiecEasi package (v.1.1.1) ^11,12^, which applies an internal centered log-ratio (CLR) transformation to account for the compositional nature of microbiome data. Taxa were filtered to those present in ≥10% of samples. Statistical significance was assessed by bootstrap permutation testing (R = 1,000 bootstraps) with BH correction, and only associations with adjusted *p* < 0.05 and |r| > 0.3 were retained. Networks were constructed as undirected graphs with node size scaled to mean relative abundance and edge width scaled to correlation magnitude. Community structure was identified using the Louvain algorithm, and hub taxa were defined as nodes with degree in the top 90th percentile. Network topology and centrality metrics were computed using igraph (v.2.1.3) ^13^ and networks were visualized using ggraph (v.2.2.1) ^14^ with a Fruchterman–Reingold layout.

**Differential abundance analysis**

Differential abundance of bacterial species across postnatal antibiotic exposure groups was assessed at each sampling time point using ANCOM-BC2, implemented in the ancombc R package (v.1.6.4) ^15^, with the “Naive” group as reference. Models were fitted separately per time point using species-level count data; taxa with zero counts and samples with zero library size were excluded prior to fitting. Both global and pairwise group comparisons were performed with pseudo-count sensitivity analysis enabled (pseudo_sens = TRUE). P-values were adjusted using the Benjamini–Hochberg method, and species with adjusted *p*-value (*p.adj*) < 0.05 were considered differentially abundant.

**Microbiota age prediction**

Microbiota age was estimated using Random Forest regression implemented in the randomForest R package (v.4.7.1.2 ^16^; ntree = 10,000, importance = TRUE), regressing MetaPhlAn4 ^17^-derived species-level relative abundances against chronological age (days of life). The model was trained exclusively on antibiotic-naive infants using a subject-level 80/20 split, ensuring no samples from the same infant appeared in both training and test sets. The optimal number of age-discriminatory taxa was identified by minimizing mean cross-validation error across 100 iterations of fivefold cross-validation (rfcv function), yielding a sparse model of 15 taxa. Feature importance was assessed using the mean decrease in mean squared error (%IncMSE) across 100 iterations. Model performance was evaluated using Spearman correlation between predicted and chronological age on held-out test samples. The finalized sparse model was subsequently applied to all samples to generate predicted microbiota age estimates. The microbiota-for-age Z-score (MAZ) was computed for each sample as the deviation of predicted microbiota age from the median predicted age of antibiotic-naive infants at the same time point, normalized by the corresponding standard deviation, as previously described ^18^. Between-group differences in MAZ at each time point were assessed using pairwise Wilcoxon rank-sum tests with BH correction.

**Community type characterization and transition dynamics**

Nasopharyngeal microbiota community types were identified using Dirichlet multinomial mixture (DMM) modeling implemented in the DirichletMultinomial package (v.1.38.0) ^19^, applied to genus-level count data filtered to genera present in ≥10% of samples. The optimal number of community types (CT) was determined by convergent minimum values across AIC, BIC, and Laplace model selection criteria, yielding k = 4 community types. Differences in alpha diversity between community types were assessed using pairwise Wilcoxon rank-sum tests with BH correction. Associations between community type distribution and time point, and between community type and antibiotic exposure group, were evaluated using Pearson's Chi-squared test. To examine microbiome stability over time, Markov chain transition probability matrices were computed separately for each antibiotic exposure group using only transitions between immediately consecutive time points within each infant, implemented in the markovchain R package (v.0.9.5) ^20^. Self-transition probabilities served as a measure of community state stability and transition networks were visualized using DiagrammeR (v.1.0.11) ^21^.

**Random Forest Classification — *S. marcescens* Persistence**

To predict long-term *S. marcescens* persistence, species-level relative abundances from samples collected at time point 1 and time point 2 were used to classify infants by *S. marcescens* colonization status at six months corrected age, defined as the presence of *S. marcescens* (relative abundance > 0) in nasopharyngeal metagenomic profiles. Random Forest models were trained using the R package randomForest (v.4.7.1.2) ^16^ via caret ^22^ with fivefold cross-validation (ntree = 500). Sparse models were constructed by selecting the top N features by across-sample variance (N = 10, 20, 30, 50) alongside a full model, with the best-performing feature set selected per time point and condition. To determine whether predictive signal was independent of *S. marcescens* abundance, models were additionally trained after excluding *S. marcescens* from the feature set. Model performance was evaluated using cross-validated accuracy, sensitivity, and specificity. Zero-variance predictors were removed before model training.
