## Supplementary figures and images for "Outbreak and Postnatal Antibiotic Exposures Drive the Development Trajectory of the Nasopharyngeal Microbiota in the First Year of Life"

### Supplementary Figures

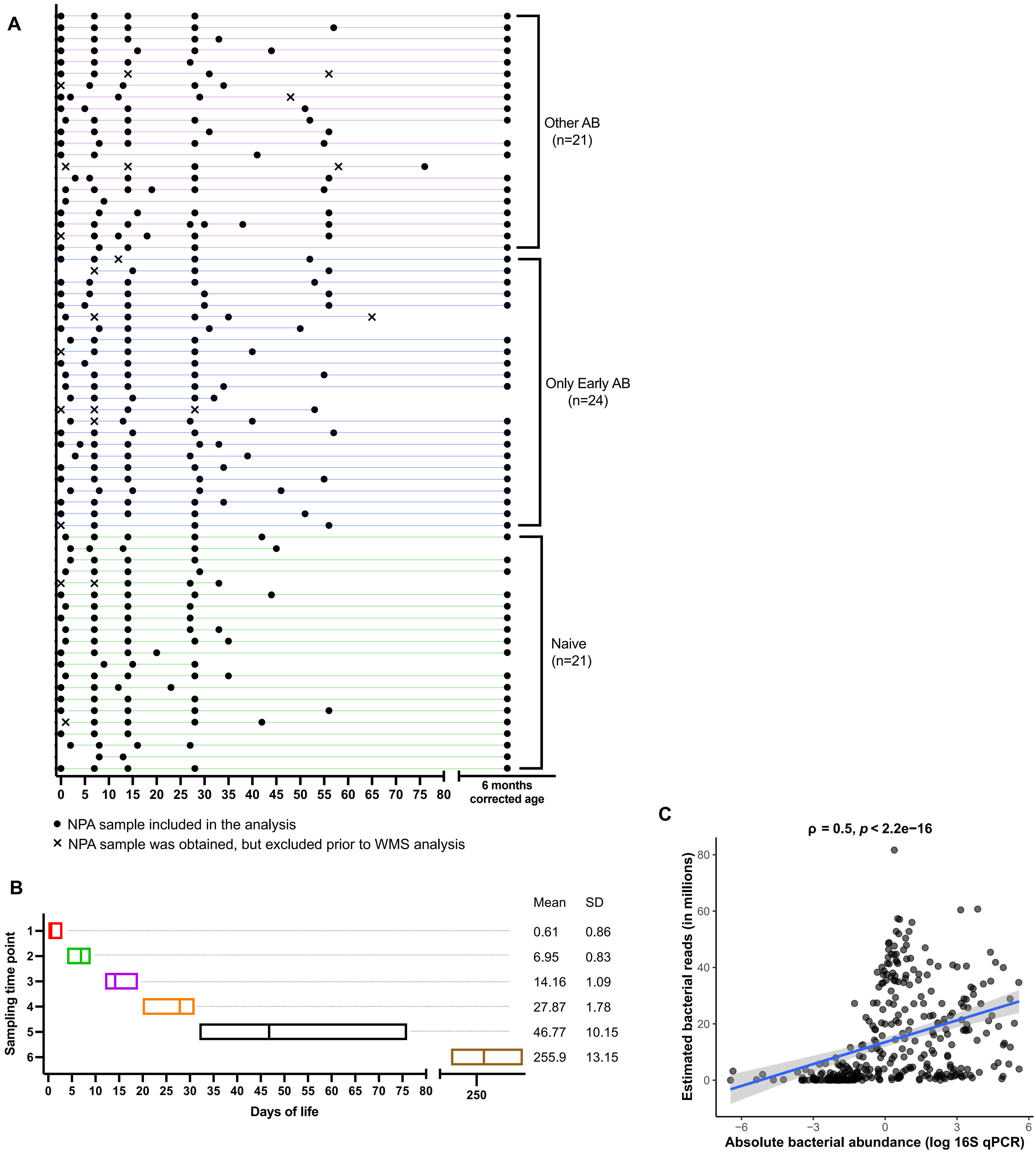

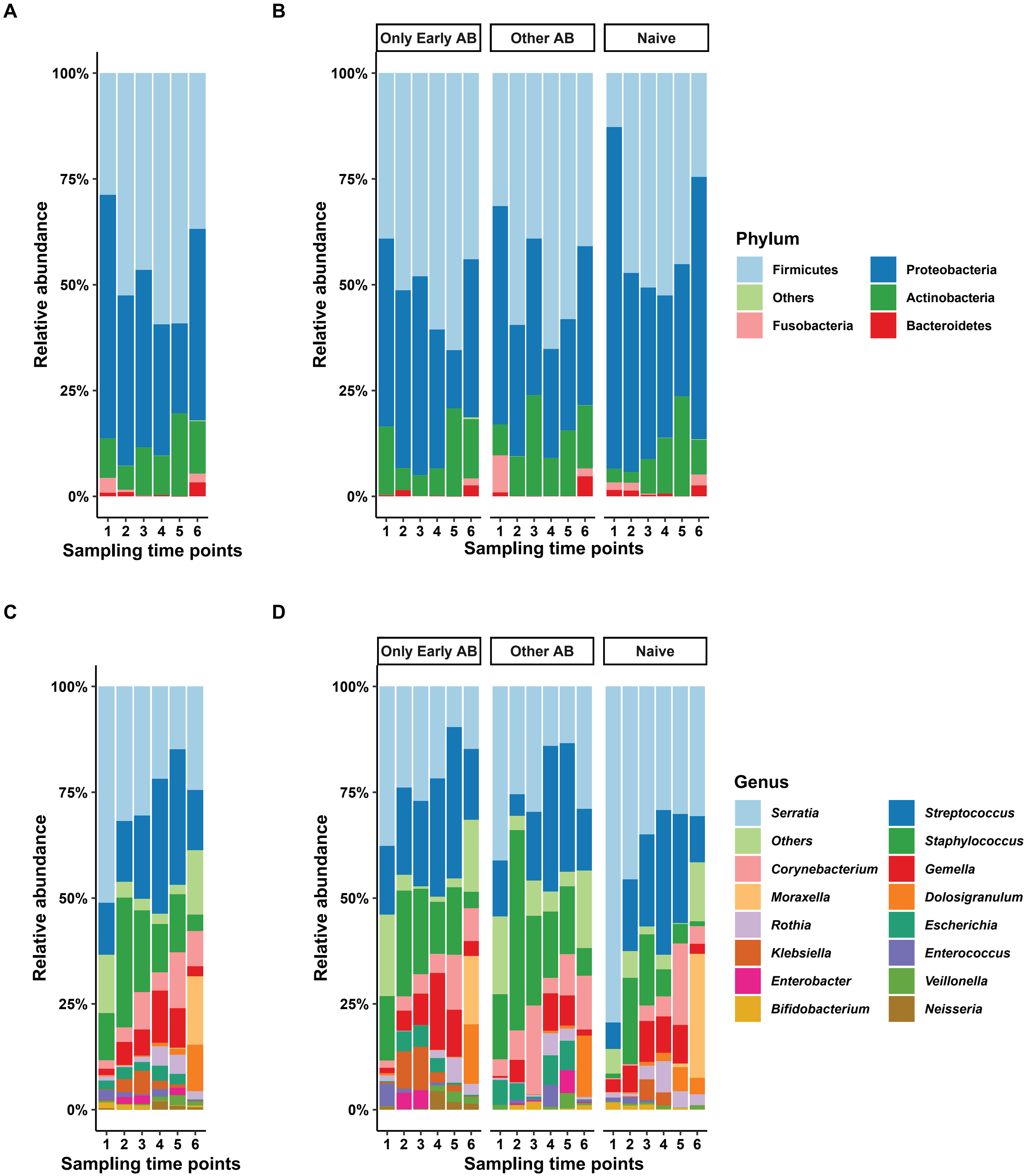

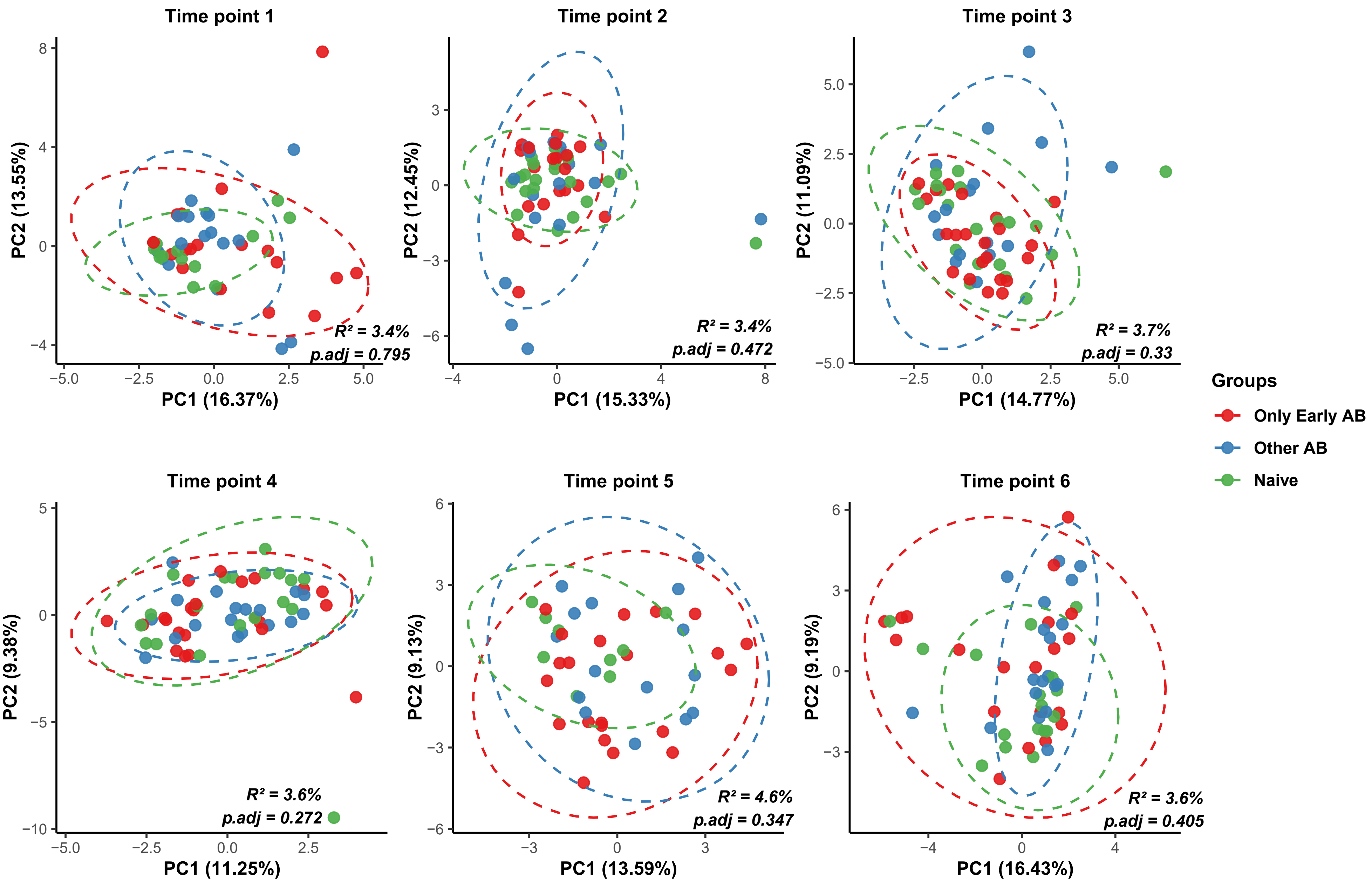

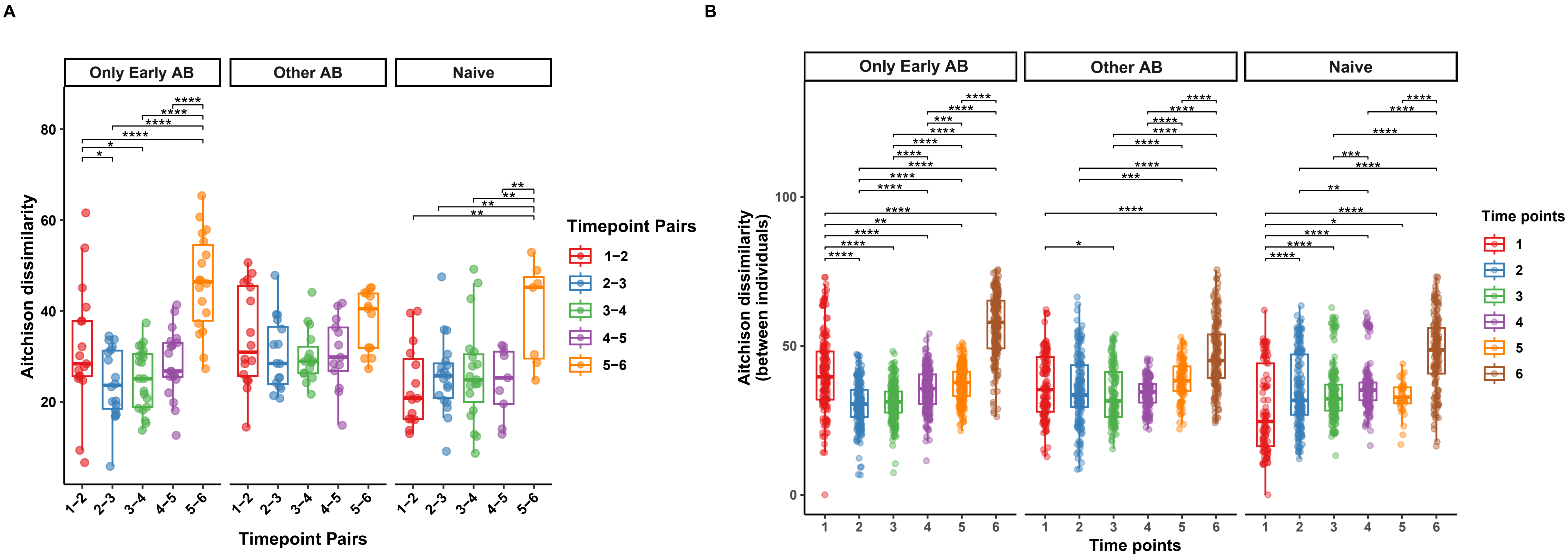

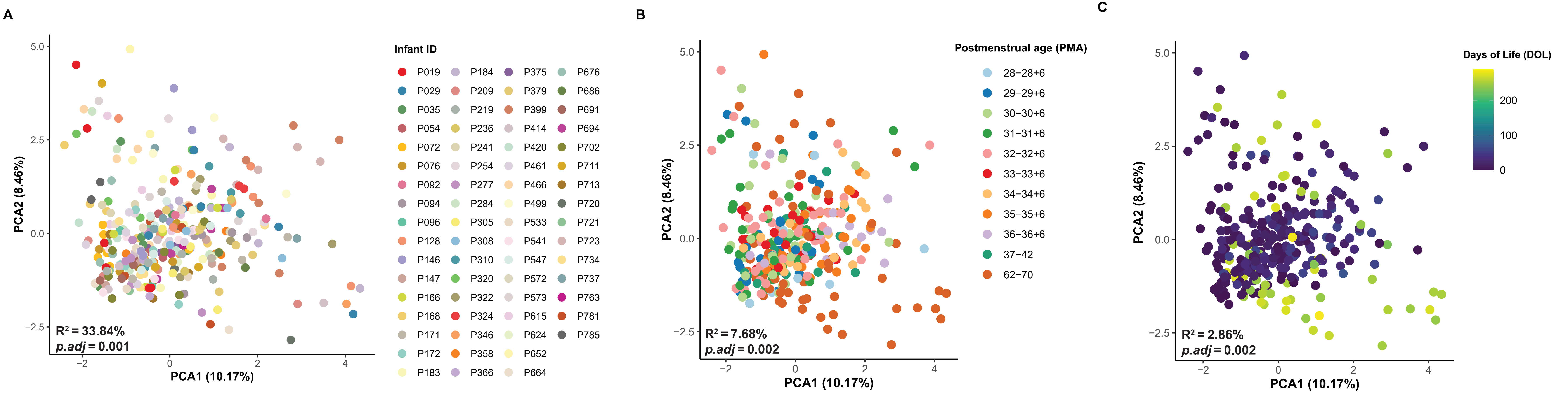

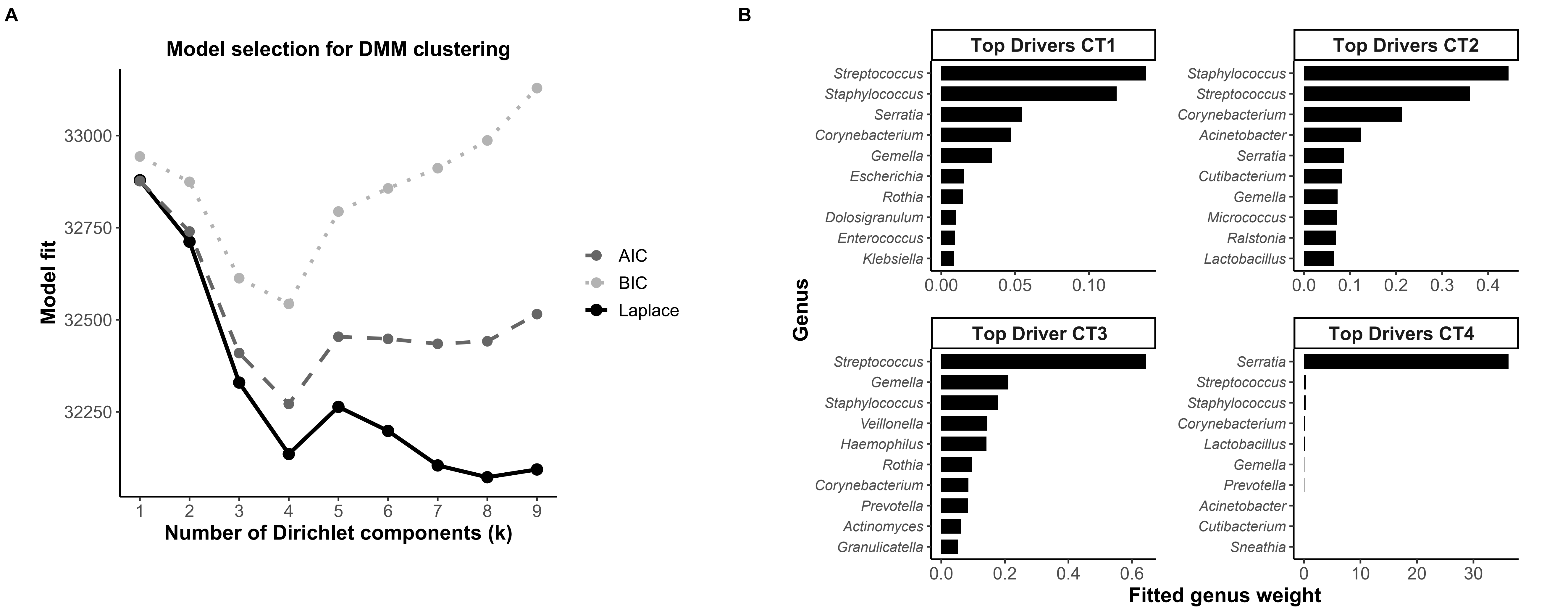

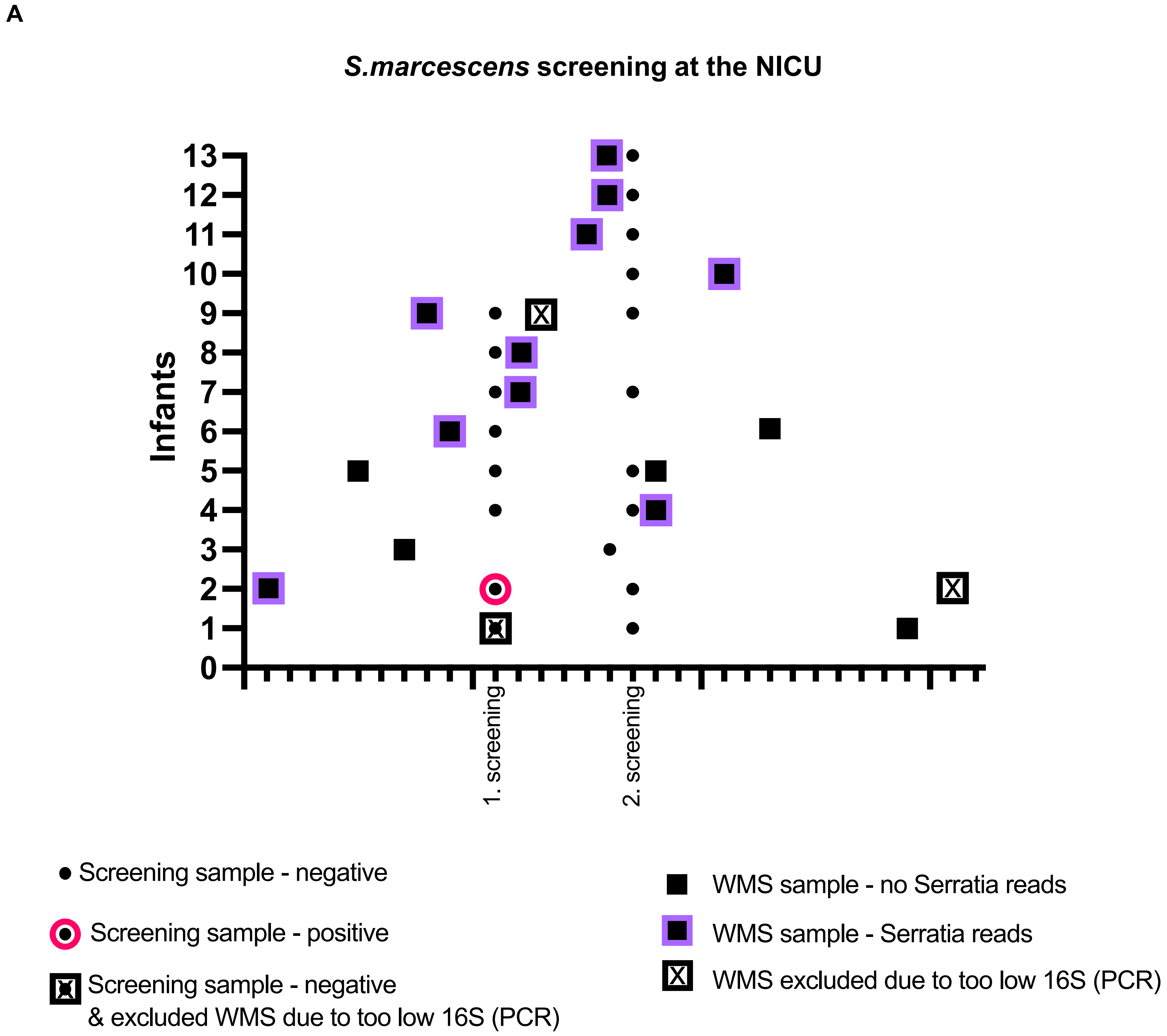


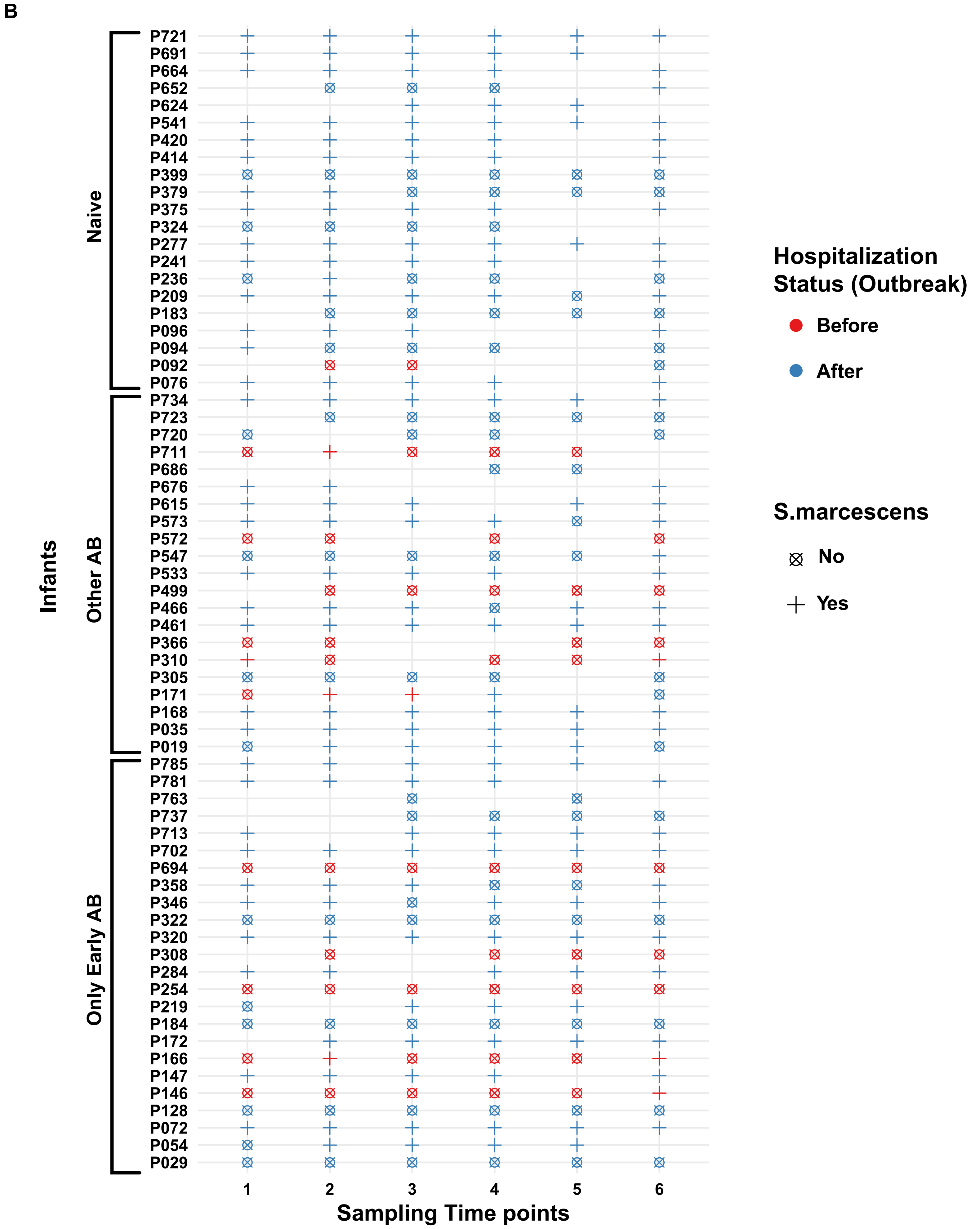


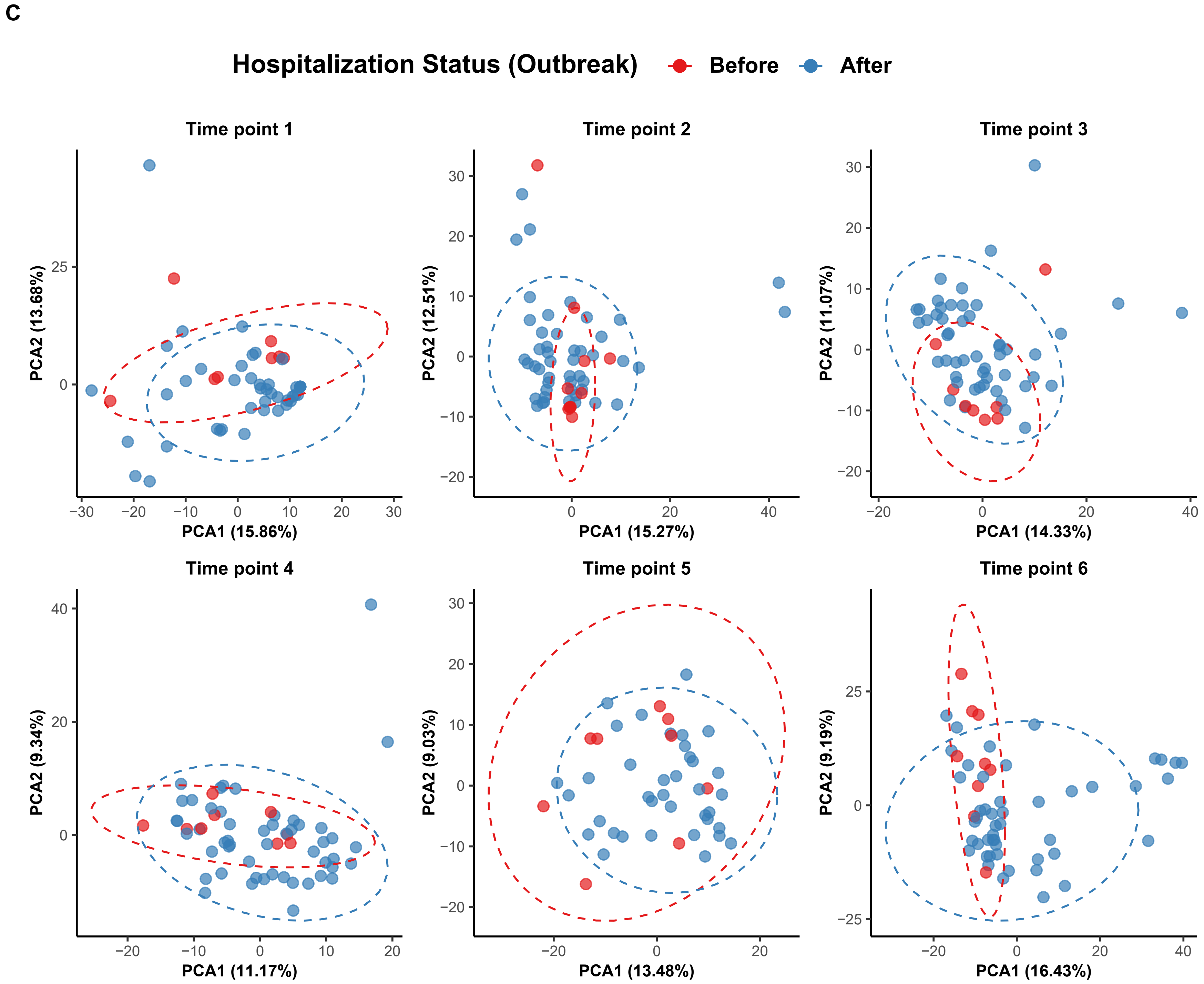


| Timepoints | $R^2$ (%) | *p.adj* |
|---|---|---|
| 1 | 4.260 | 0.039 |
| 2 | 3.117 | 0.048 |
| 3 | 2.714 | 0.073 |
| 4 | 2.725 | 0.048 |
| 5 | 3.087 | 0.088 |
| 6 | 3.753 | 0.039 |

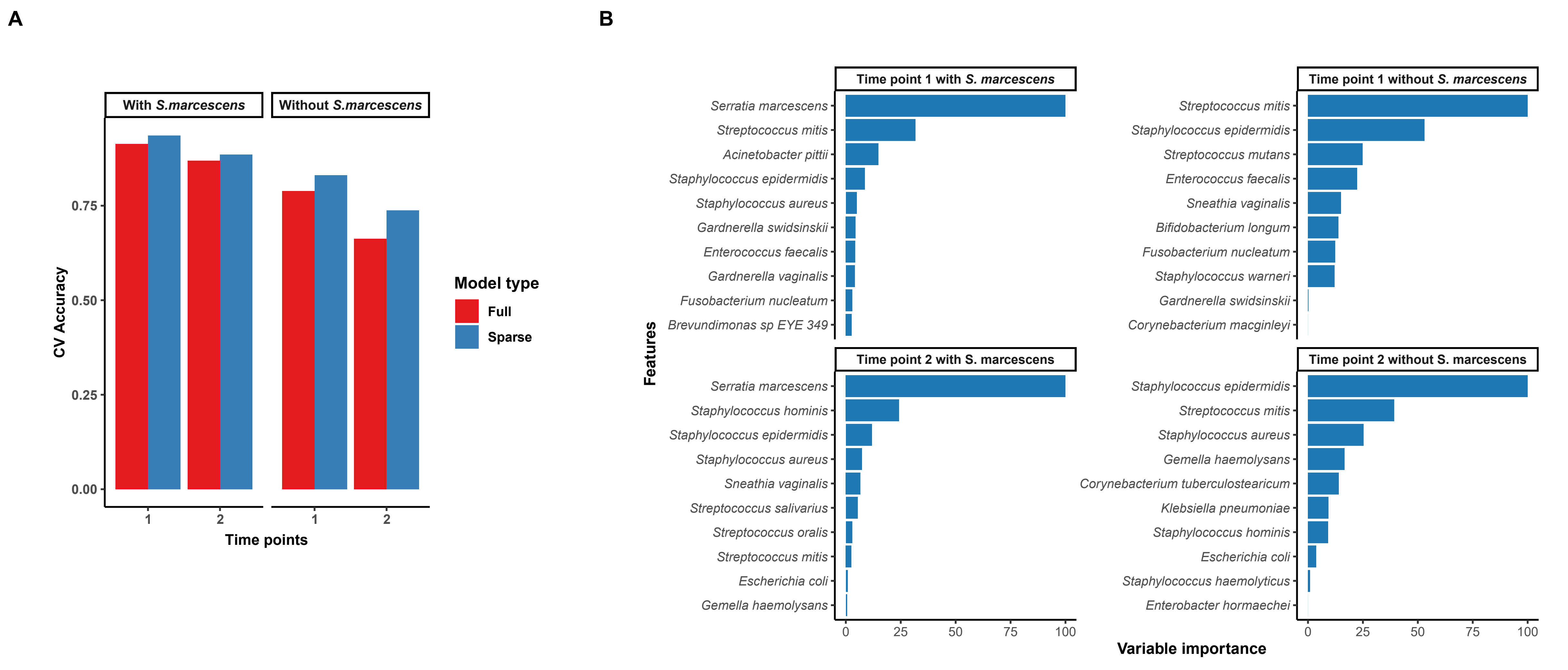

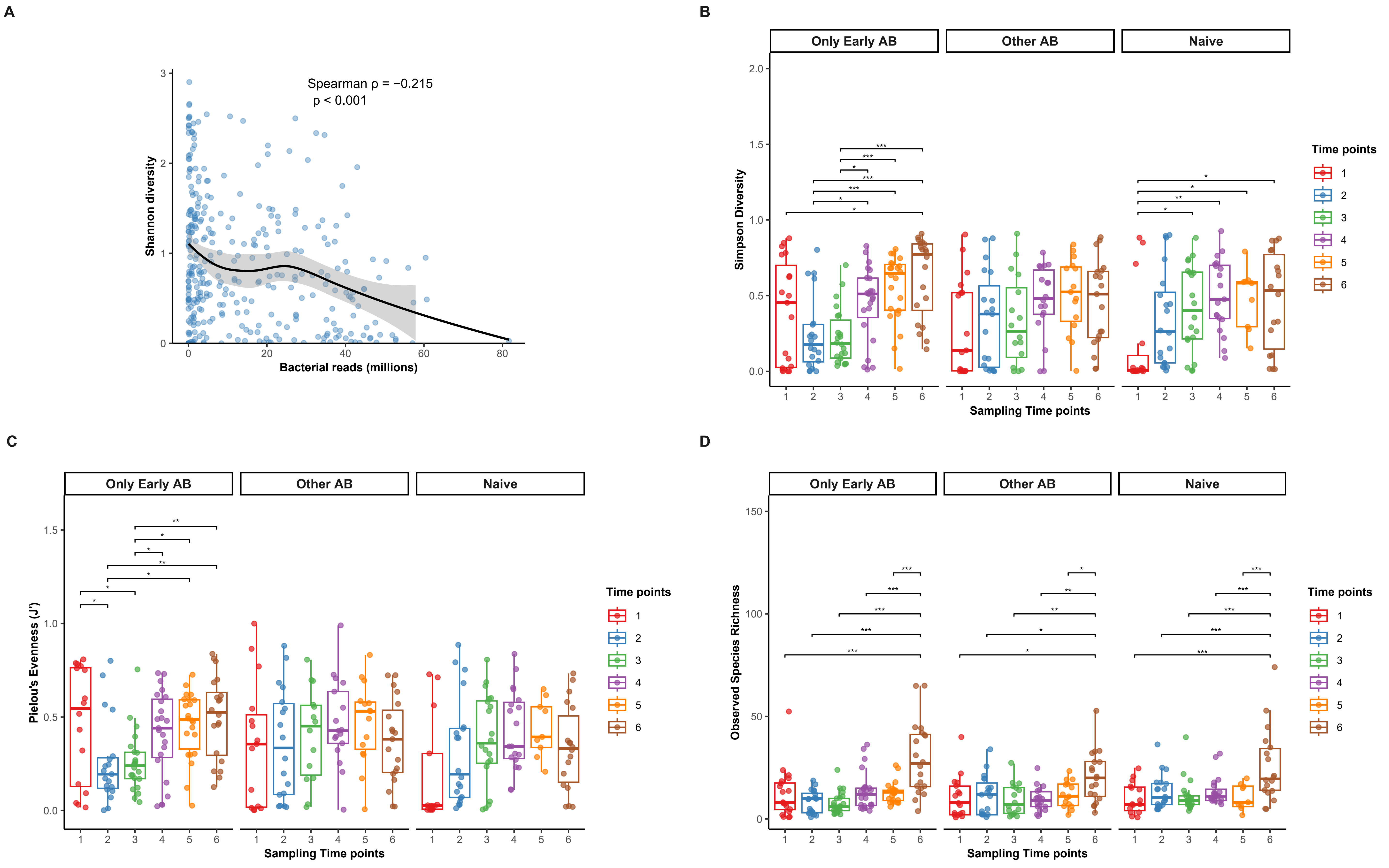
